# Effects of nationwide alerting for acute kidney injury on healthcare and patient outcomes: population based, regression discontinuity analysis

**DOI:** 10.1101/2025.10.02.25337043

**Authors:** Min Xie, Pascal Geldsetzer, Tom Blakeman, Matthew James, Tim Scale, Zhi Tan, Simon Sawhney

## Abstract

**Objective:** To determine whether the implementation of electronic alerts for acute kidney injury improves care and outcomes in real-world clinical practice.

**Design:** Population-based, regression discontinuity analysis

**Setting:** Hospital and community-based systems across all 8 health boards in Wales, from 2016 to 2021 following implementation of AKI alerts.

**Participants:** 3.1 million adults aged ≥18 years resident and registered at GP practices in Wales between the years of 2016-2021, following implementation of AKI e-alerts across all 8 Wales health boards.

**Exposure:** Electronic alerts for acute kidney injury provided in either a passive (7 boards) or interruptive (1 board) manner to physicians through laboratory reporting systems.

**Main outcome measures:** Mortality, hospital admission or readmission, severity and recovery of AKI, coding of acute kidney injury, proteinuria and blood pressure measurements.

**Results:** Among 861,494 and 354,505 eligible patient encounters, 5.8% and 2.0% AKI alerts were generated from hospital and community settings, respectively; mean age 64 years, 54% female. In both hospital and community settings, respectively, AKI alerts led to no significant differences in mortality [complier average treatment effect +1.31% (95% CI −3.07, 4.74); +2.07% (95% CI −3.44, 6.65)] or admissions/readmissions [+0.13% (95% CI −3.82, 4.21); +4.07% (95% CI −1.84, 8.27)]. There was a modest increase in hospital coding of AKI with alerts [+5.88% (95% CI 2.22, 7.58)], but no difference in primary care coding of AKI after discharge [+0.72% (95% CI −0.67, 1.30)]. Alerts exerted small increases on subsequent checks for proteinuria and of blood pressure. Findings were consistent for passive and interruptive alerts. There were no meaningful differences by rurality, deprivation, sex, history of surgery, diabetes, or vascular disease.

**Conclusions:** The nationwide implementation of AKI alerts in Wales produced small effects on documentation of AKI and some processes of care but exerted no effects on survival or hospital admissions in acute care or community settings. The consistently poor outcomes, and the deficiencies in documentation and care after AKI highlight the ongoing need for an improved clinical response.

**Registration:** A prespecified analysis protocol is available at Open Science Framework (OSF) repository (https://osf.io/f4wz9/?view_only=d20e49817bc74f1fa66afc7aa8dda0ab)

**Summary:** *What is known:* Previous randomised trials and real-world observational studies of electronic alerts for acute kidney injury (AKI) have produced mixed results. The applicability of previous trials for real-world clinical practice remains uncertain and contested. AKI e-alerts are still widely used.

*What we did:* We applied Regression Discontinuity Design (RDD), a method gaining traction in clinical research. RDD can work like an ideal target trial to enable causal inference in real-world settings. RDD is particularly effective when clinical actions are triggered by a specific measurement threshold. This approach allowed us we to evaluate a nationwide AKI e-alert initiative across Wales.

*What we found:* We found no evidence that AKI e-alerts improved or worsened outcomes across any clinical setting, patient subgroup, or alert delivery method. Nonetheless, the consistently poor outcomes, and the deficiencies in documentation and care after AKI highlight the ongoing need for an improved clinical response.

## Introduction

Use of electronic health records to deliver care is growing^1^. Electronic alerts (e-alerts) are one form of decision support used in electronic systems to draw the attention of health professionals to the presence of important clinical abnormalities. One example where e-alerts have been broadly leveraged is for identification of acute kidney injury (AKI), an abrupt loss of kidney function typically detected by an increase in serum creatinine. AKI is common, associated with adverse short- and long-term outcomes, and opportunities to improve care have been identified in past audits.^9, 10^ In 2014, NHS England published a major patient safety directive mandating the implementation of a standardised serum creatinine-based early warning algorithm for AKI into Laboratory Information Management Systems across England,intended to improve recognition and response^11^. Between March 2014 – March 2015, a version of this algorithm was introduced across Wales in both primary and secondary care and reported as an “AKI alert” in patient records. This initiative was a key element of NHS England’s Think Kidneys Programme, which entailed six multidisciplinary workstreams^12^ to prevent avoidable harm caused by AKI with tools and resources for patients and professionals to support the prevention, early detection, management of AKI across systems of care^13^.

AKI alerts continue to be used widely, but evidence for their benefit has been conflicting. Proponents of AKI alert systems point to large observational and “before-and-after” real-world studies^14, 15^ which have reported small but meaningful decreases in hospital mortality, dialysis use, kidney recovery^16–23^, and a cluster randomised trial that reported reduced hospital stay as part of a multifaceted intervention^24^. However, two randomised clinical trials of AKI alerts delivered by phone or pager in hospital settings in the United States failed to show improvements in hospital mortality or kidney outcomes, with potential harm in some subgroups^25, 26^. Trials of e-alerts are limited by potential contamination between intervention and control groups, uncertainty about generalizability beyond trial settings, and limited power to detect small intervention effects. Observational real-world studies are limited by confounding effects from imbalances in case-mix between intervention and control groups; coincidental hospital or system level changes that may introduce other imbalances between alert and non-alert populations; and selection biases arising out of shifts over time in underlying patterns of testing leading to increases in the detection of AKI cases. Thus, the effectiveness of creatinine threshold-based AKI alerts remains uncertain despite continued delivery.

To address the discrepancies in evidence from trials and observational studies of AKI alerts, we used a regression discontinuity design (RDD) to examine the effects of implementation of AKI alerts in Wales on processes of care and clinical outcomes in both hospital and community settings. The nature of implementation of AKI alerts in Wales enabled us to further examine the effects of both passive and interruptive delivery.

## Methods

### Study Design

The arbitrary nature of creatinine thresholds for the definition of AKI (a rise of 26 micromol/L within 48 hours or 50% increase within 7days) creates an opportunity for a natural experiment that can be analysed using regression discontinuity.”^27, 28^ In this study, we focused on the threshold 50% creatinine rise that triggers the majority of AKI alerts in electronic systems. Given the inherent random measurement variability in serum creatinine, people with a 49% and 51% rise are expected to be identical in all characteristics except for their probability of receiving an AKI alert. Under this assumption, patients close to but below and above the threshold for an AKI alert may be considered to be effectively randomized to receive an AKI alert versus no alert at the time of creatinine measurement (pseudorandomisation), equivalent to an ideal target trial.

### Study Population

Data for the study were obtained from the SAIL databank of linked routine health records of hospital, primary care, and all laboratory data (including serum creatinine results and AKI alerts)^29–33^. The study population included 3.1 million adults aged ≥18 years resident and registered at GP practices in Wales between the years of 2016-2021, following the implementation of AKI e-alerts across all 8 Wales health boards. People were included in the study only if they had at least two serum creatinine tests conducted within a period of 365 days, a baseline estimated glomerular filtration rate (eGFR) using the 2009 race-free CKD EPI formula^34^ of ≥15 ml/min/1.73m^2^ and did not have a documented history of long-term dialysis, kidney transplant or kidney failure (chronic kidney disease stage G5).

Two distinct study populations were created to assess the impact of AKI alerts in two different contexts (figure S1):

1. Hospital subpopulation: People admitted to hospital as an emergency across 18 acute hospitals in Wales, who had no evidence of AKI in the 7 days prior to hospitalisation. Analysis (pseudo-randomisation) was based on the first serum creatinine test taken during the admission. In a sensitivity analysis we also broadened the definition to use the highest serum creatinine during the first 2 days of hospital admission as the qualifying creatinine test for pseudorandomisation.
2. Community subpopulation: People in the community who have recently been discharged from hospital have a particularly high risk of AKI and readmission. Thus, among people discharged following an emergency hospital admission without AKI, the analysis was based on the first of any community serum creatinine taken within 90 days of discharge.

### Baseline covariates

Variables used to evaluate baseline balance between patients below and above the alert triggering threshold included age, sex, Welsh index of multiple deprivation (WIMD), rural resident neighbourhood, baseline eGFR, previous history of coronary disease, peripheral arterial disease, stroke, heart failure, diabetes mellitus, and prescription of a statin, proton pump inhibitor (PPI), nonsteroidal anti-inflammatory drug (NSAID), loop diuretic, and renin aldosterone angiotensin system inhibitor (RAASI) in the prior 180 days.

### Exposure

The exposure (AKI alert) was defined as having a record of an AKI alert delivered on the day of the serum creatinine test of interest. For the evaluation of the effect of alerts in the hospital setting, we analysed the effects of interruptive and passive alerts separately. Passive AKI alerts were implemented in 7 of the 8 health boards in Wales and appeared as a standardised text underneath the creatinine values with a hyperlink to the Welsh AKI guidelines. An interruptive alert was implemented in one health board (Cwm Taf) and involved the addition of a telephone call from the laboratory to a member of the patient’s clinical team who were directed to complete and record a care bundle (figure S2), and with notification of a hospital outreach service. All stages of AKI were phoned through to primary and secondary care, excluding intensive care and dialysis units (figure S3).

### Outcomes

#### Main outcomes

Prespecified outcomes included both processes of care (changes in clinical documentation and care), and clinically important patient health outcomes. The two co-primary clinical outcomes were mortality within 1 year and non-elective hospital admission/readmission or emergency department attendance within 180 days. We also evaluated mortality and admission/readmission within 30 days.

#### Secondary outcomes

Additional secondary clinical outcomes included AKI severity determined by the peak creatinine level during hospitalisation (for the hospital subpopulation), and kidney function based on the updated eGFR at 90 days (among those with an eGFR within 90 days). Care process outcomes were prolonged hospital length of stay of longer than 7 days^24^, post-discharge blood pressure and creatinine test within 90 days, proteinuria assessment within 1 year (urine albumin/creatinine ratio, protein/creatinine ratio, or dipstick urinalysis), and coding for AKI in hospital or within 90 days in primary care. Medication outcomes were subsequent prescriptions for oral NSAIDs, PPIs, continuation of loop diuretics (among those with a prescription dispensed in the previous 180 days), and continuation of RAASIs (including angiotensin converting enzyme inhibitors and angiotensin receptor blockers among those with a prescription dispensed in the previous 180 days).

Negative control outcomes, for which no effect of AKI alerts would be expected, were new diagnosis of cancer within 1 year, and continuation of a statin among those prescribed in the previous 180 days. For secondary outcomes and negative control outcomes, the main analyses were restricted to patients who survived to the end of the ascertainment period. Except for mortality, peak creatinine during hospitalisation and length of stay, the outcomes were followed up from discharge from the indexed hospitalisation for the hospital setting, and from the indexed creatinine test for the community setting.

### Statistical analyses

The RDD employed in this study relied on a specific threshold rule, which triggers an AKI alert when the relative increase in serum creatinine exceeds 50%. Based on this threshold rule, we defined a centred running variable as the absolute change in serum creatinine relative to 1.5 times the baseline reading (in micromol/L). An alert would be triggered when the centred running variable exceeds 0 micromol/L. Imperfect compliance with this threshold rule could arise due to alternative alert-triggering rules (e.g. absolute creatinine rise of 0.3mg/dL, 26 micromol/L, within 48 hours) or measurement error in electronic health records (EHRs). In such cases, the observed discontinuity at the threshold provides an unbiased estimate of the effect of the threshold rule, also known as the intent-to-treat (ITT) effect. By scaling the ITT effect by the discontinuity in the probability of triggering an alert, we estimated the complier average treatment effect (CACE) - the effect of alerting among compliers. This derivation, known as “fuzzy RDD”, is essentially an instrumental variable approach^35^. Following the standard practice, we used local linear regressions with triangular weights, restricting the analyses to observations within a narrow window around the threshold (“bandwidth”). The bandwidths were chosen to be mean-squared-error (MSE) optimal^36^. Asymmetry in the running variable’s distribution was accounted for by allowing different bandwidths on either side of the threshold. We calculated bias-adjusted robust confidence intervals (CIs) to ensure reliable inference of the effect estimates^37^. Additional details on the RDD for this analysis are in the supplementary material.

We reported the CACE for each outcome. Additional analyses were carried out to assess the robustness of estimates to alternative bandwidth choices, functional forms, and the inclusion of patients who died before follow-up completion.

To address potential confounding and biases, we conducted two falsification tests following: (1) applying the analysis to negative outcome controls, for which no effect should be expected^38^, and (2) analysing the main outcomes in data from January 1, 2010, to January 1, 2014, prior to the implementation of AKI alerts. In this pre-alert period, no threshold effect would be anticipated, ruling out confounding effects on AKI alerting.

Finally, subgroup analyses explored effect heterogeneity across predefined groups, including individuals admitted to hospitals that implemented interruptive and passive AKI alerting systems, from the most deprived quintile of the Wales Index of Multiple Deprivation (WIMD), by sex, recent major surgery, history of vascular disease, and history of diabetes mellitus. These represent subpopulations in whom AKI care and outcomes might be expected to differ or respond differently to alerts.

All RDD analyses were performed using the *rdrobust* package (v.2.2) in R (version 4.3.3).

Ethics statement: This study was conducted within the Secure Anonymised Information Linkage (SAIL) Databank. Approval for the use of anonymised data in this study was obtained from the SAIL independent Information Governance Review Panel (IGRP; application number 1306).

Patient/public involvement: The Welsh AKI initiative was based on a contemporaneous patient safety alert from NHS England, and included multi-stakeholder engagement from the Welsh AKI Steering Group, however the steering group did not include patients or public involvement at the time of implementation. We are grateful to members of the Grampian Kidney Patient Association who helped us to think through how we could effectively and acceptably use routine health data to help inform changes in AKI care.

## Results

### Population Characteristics

During 2016-2021, there were 861,494 and 354,505 hospital and community presentations eligible for inclusion, including 5.8% and 2.0% that triggered an AKI alert, respectively. Mean ages were 64.6 and 64.2 years, and 54.5 and 54.4% of people were female. Across hospital and community settings a minority had a history of CKD (29.3%, 24.7%), diabetes (23.6%, 24.7%), heart failure (16.7%, 16.3%), RAASI use (28.0%, 31.2%) and loop diuretic use (17.0%, 18.5%) (Table S1).

### Satisfaction of RDD assumption

In each setting (hospital and community), we assessed the fundamental assumption of RDD, which posits that patients just below and just above the threshold are exchangeable. This assumption would be compromised if patients or physicians could intentionally manipulate creatinine test results to place patients on a specific side of the threshold, either to avoid or to trigger an alert. Such manipulation would not be expected in this study as precise creatinine test results are directly generated and uploaded by the laboratory into the electronic health data system, eliminating any potential for interference by either patients or physicians. This was supported by two sets of findings. First, there was no evidence of heaping in the distribution of the centered running variable on either side of the threshold (figure S4). Second, no discontinuities were visually detected at the threshold in key baseline characteristics, including age, baseline estimated glomerular filtration rate (eGFR), and WIMD deprivation decile, further indicating that these variables did not act as confounders (figure 1). These results were corroborated by applying RDD to these and other baseline characteristics, empirically demonstrating that all relevant baseline variables were continuously distributed and balanced near the threshold (table S2). In contrast, a significant discontinuity was observed at the threshold in the probability of an AKI alert being triggered in both settings (figure 1). Collectively, these analyses support the validity of the RDD assumptions, confirming that the threshold effectively differentiates alert delivery while maintaining the exchangeability between groups.

**Figure 1.**
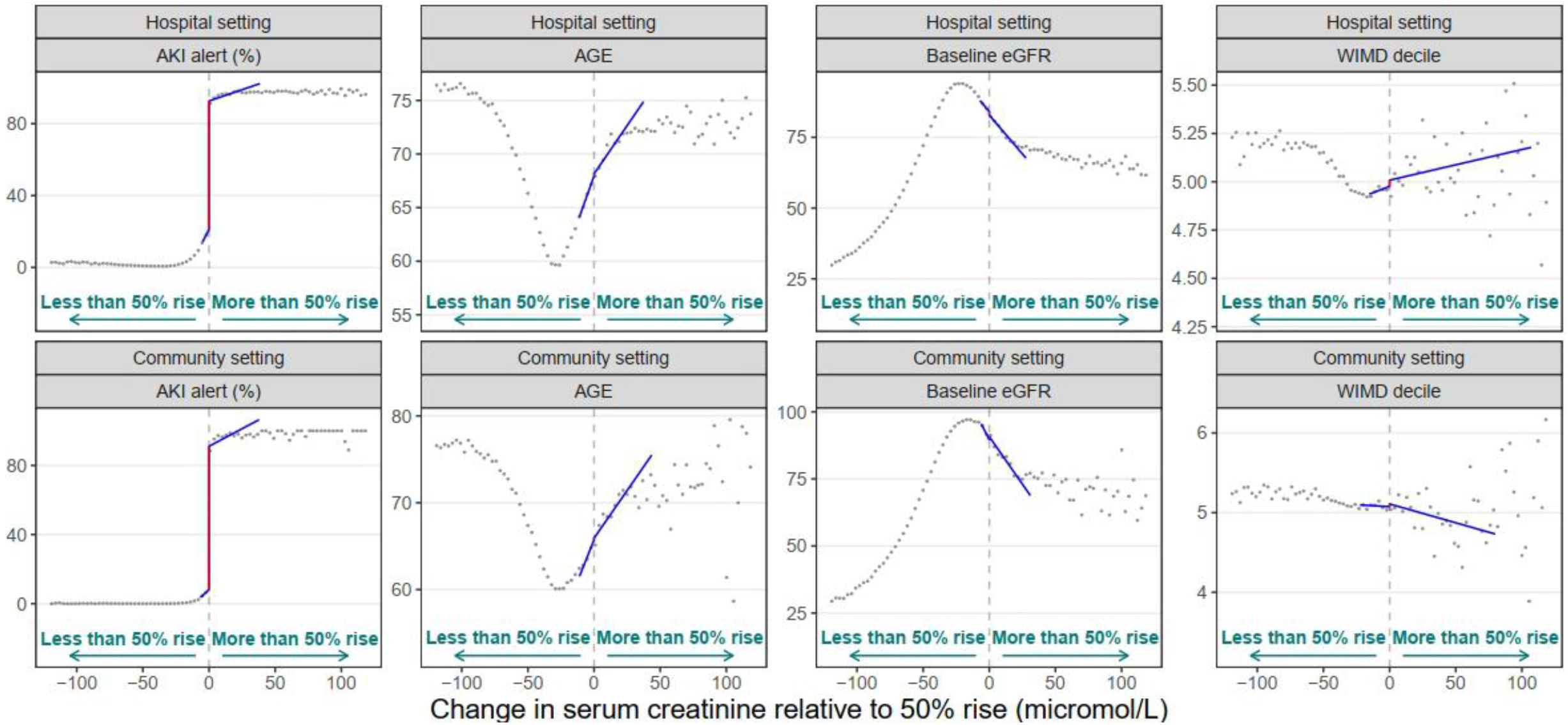
Association between the change in creatinine and AKI alert delivery and potential confounders. ^**1**,**2**,**3**^. ^1^ The data source for this analysis was the SAIL databank. ^2^ Grey dots show the mean value for each bin of change in creatinine. Red vertical lines denote the discontinuity. ^3^ The blue lines are fitted by our main RDD model. Abbreviations: AKI = acute kidney injury; eGFR = estimated glomerular filtration rate; WIMD = Welsh index of multiple deprivation *Notes*: At the threshold for delivery of AKI alerts there was a large discontinuity in the likelihood of an alert being delivered with no difference (i.e. baseline balance) in age, baseline eGFR, and deprivation measure. These findings support the RDD assumption that patients above and below the threshold differed only in receipt of an AKI alert and not in other baseline characteristics

### Effect of AKI alerts on the primary outcomes; mortality and admission

Table 1 summarises the estimated effects of AKI alert (i.e. CACE) on the coprimary clinical outcomes, secondary kidney outcomes, and clinical care processes for hospital and community settings. Irrespective of setting, mortality was substantial among individuals just below the 50% creatinine rise threshold (29.81% and 25.32% at 365d, and 11.66% and 6.64% at 30d for hospital and community settings respectively). Mortality was unaffected by AKI alerts, with absolute differences in mortality of +1.31% (95% CI −3.07, 4.74) in the hospital setting and +2.07% (95% CI −3.44, 6.65) in the community setting.

**Table 1.** Effect of AKI alert^1,2,3^. ^1^Mean below threshold is calculated as the mean within the 5 micromol/L below the threshold. ^2^For each outcome, the MSE-optimal bandwidth is included in Table S7 in the supplement. ^3^Effective sample size is the number of observations within the MSE-optimal bandwidth. *Notes*: A positive estimate indicates the AKI alert associated with an increase in the outcome in the units of the outcome (%, unless otherwise stated)

|  | Hospital setting |  |  |  | Community setting |  |  |  |
| --- | --- | --- | --- | --- | --- | --- | --- | --- |
|  | Mean below threshold | Effect of AKI alert |  | Effective sample size | Mean below threshold | Effect of AKI alert |  | Effective sample size |
|  |  | Estimate (95% CI) | P value |  |  | Estimate (95% CI) | P value |  |
| **Primary outcome** |  |  |  |  |  |  |  |  |
| Death in 365 days (%) | 29.81 | 1.31 (-3.07, 4.74) | 0.674 | 34,685 | 25.32 | 2.07 (-3.44, 6.65) | 0.534 | 9,590 |
| Emergency hospitalization or emergency department attendance in 180 days (%) | 52.98 | 0.13 (-3.82, 4.21) | 0.924 | 33,793 | 61.18 | 4.07 (-1.84, 8.27) | 0.212 | 10,592 |
| **Secondary outcome** |  |  |  |  |  |  |  |  |
| *Heath outcome* |  |  |  |  |  |  |  |  |
| Death in 30 days (%) | 11.66 | 0.43 (-2.33, 2.52) | 0.941 | 37,112 | 6.64 | 0.58 (-2.02, 2.30) | 0.9 | 17,598 |
| Emergency hospitalization or emergency department attendance in 30 days (%) | 26.00 | -0.35 (-3.33, 2.54) | 0.791 | 39,261 | 40.15 | 2.98 (-3.11, 7.12) | 0.442 | 11,284 |
| *Kidney recovery outcome* |  |  |  |  |  |  |  |  |
| Peak inpatient creatinine after test (micromol/L) | 106.80 | 6.45 (-0.46, 11.16) | 0.071 | 33,993 |  |  |  |  |
| Updated eGFR at 90 days (ml/min/1.73m<sup>2</sup>) | 80.04 | -0.40 (-2.75, 2.55) | 0.941 | 23,648 | 81.51 | 0.80 (-2.48, 5.30) | 0.479 | 6,130 |
| *Care process outcome* |  |  |  |  |  |  |  |  |
| Length of inpatient stay >=7 days (%) | 47.21 | -3.09 (-8.12, 0.78) | 0.106 | 32,779 |  |  |  |  |
| Having an inpatient creatinine retest (%) | 70.95 | 0.80 (-3.80, 4.21) | 0.919 | 31,945 |  |  |  |  |
| Proteinuria check in 365 days (%) | 21.84 | 4.33 ( 0.76, 7.28) | 0.016* | 28,551 | 21.50 | 1.15 (-3.76, 5.03) | 0.777 | 10,417 |
| Creatinine test in 90 days (%) | 57.98 | 1.71 (-3.12, 5.54) | 0.584 | 28,577 | 66.29 | 3.59 (-2.67, 8.05) | 0.326 | 8,788 |
| Blood pressure measurement in 90 days (%) | 35.38 | 3.79 ( 0.58, 6.79) | 0.020* | 37,947 | 36.56 | -0.56 (-5.63, 3.42) | 0.633 | 12,225 |
| AKI coding in primary care in 90 days (%) | 3.30 | 0.72 (-0.67, 1.30) | 0.527 | 72,088 | 1.97 | 3.27 ( 1.56, 4.25) | <0.001*** | 61,535 |
| AKI coding for hospitalization (%) | 16.08 | 5.88 ( 2.22, 7.58) | <0.001*** | 36,152 |  |  |  |  |
| Any PPI prescription in 365 days (%) | 44.95 | 2.10 (-1.65, 5.43) | 0.295 | 32,021 | 47.80 | -1.52 (-6.28, 3.07) | 0.501 | 11,762 |
| Any NSAID prescription in 365 days (%) | 9.91 | 0.23 (-1.73, 2.53) | 0.712 | 29,016 | 8.97 | 3.19 ( 0.57, 6.56) | 0.020* | 9,215 |
| RAASI continuation in 180 days among prior users (%) | 80.93 | 2.29 (-2.09, 7.39) | 0.274 | 11,913 | 85.90 | -2.71 (-8.74, 3.39) | 0.387 | 3,779 |
| Diuretic continuation in 180 days among prior users (%) | 79.72 | 5.07 ( 0.01, 10.18) | 0.050* | 9,286 | 82.99 | 1.76 (-4.94, 6.73) | 0.764 | 5,457 |

Similarly, while emergency hospital readmission/admission or emergency department attendances were common (52.98% and 61.18% at 180d, and 26.00% and 40.15% at 30d for hospital and community settings respectively), there was no effect of AKI alerts in either setting (increase of +0.13%, 95% CI −3.82, 4.21 for hospital setting; +4.07%, 95% CI −1.84, 8.27 for community setting). Figure 2 depicts the association between the change in creatinine (i.e. the effect of the cut off) and the two long-term health outcomes, where the estimated discontinuities at the cut off represented the ITTs.

**Figure 2.**
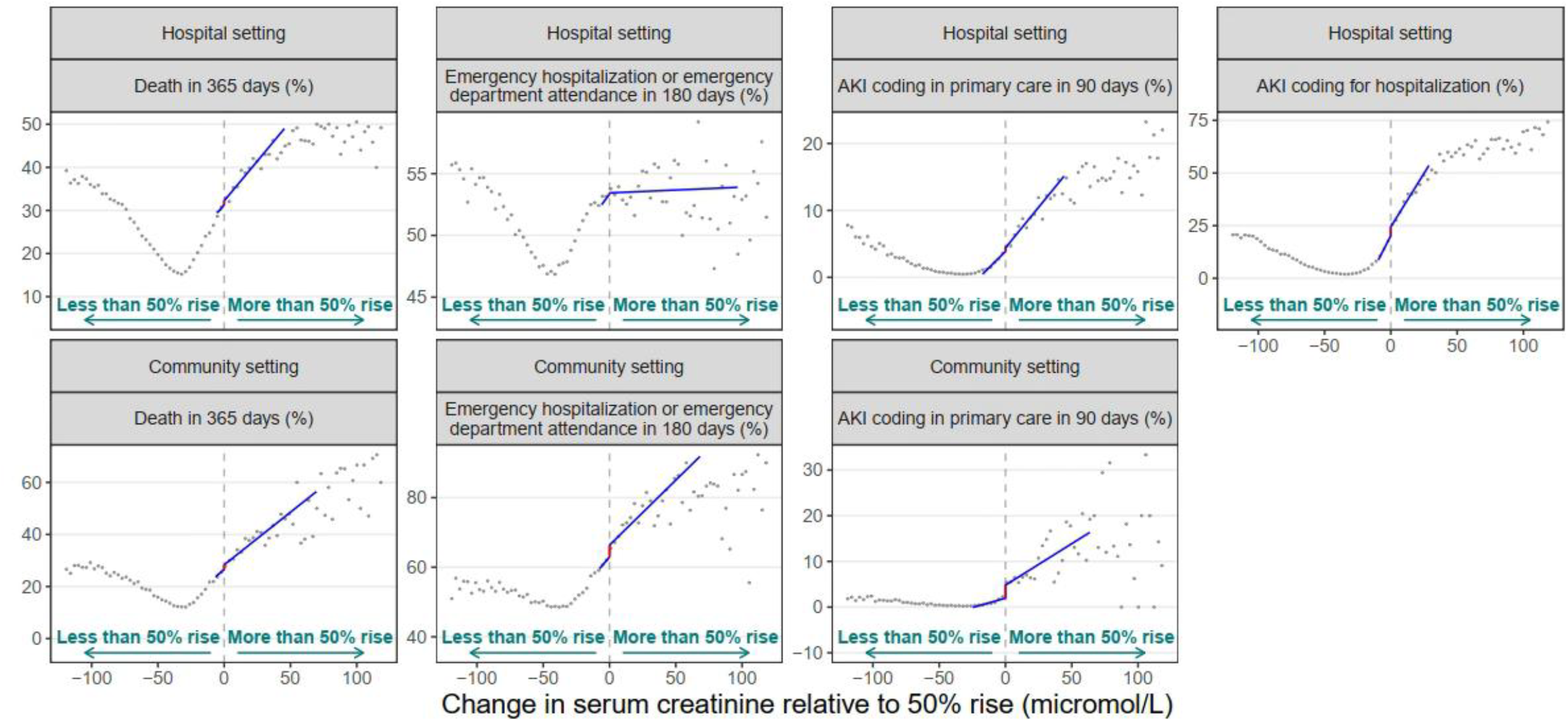
Association between the change in creatinine and outcomes ^**1**,**2**^. ^1^ Grey dots show the mean value for each bin of change in creatinine. ^2^ The blue lines are fitted by our main RDD model. Red vertical lines denote the discontinuity. *Notes*: Where AKI alerts where delivered in a hospital settings there was no significant discontinuity at the threshold for alerting in mortality, readmission, coding of AKI in primary care, and a modest increase in coding of AKI for hospitalization. In the community setting there was no significant discontinuity in mortality, and modest increases in likelihood of hospital admission, and coding of AKI in primary care.

### Effect of AKI alerts on secondary outcomes: kidney outcomes and care process

In the hospital setting, AKI alerts had no effect on repeat blood test, peak creatinine during admission, eGFR at 90 days, or likelihood of an admission prolonged beyond 7 days (table 1).

There was an increase in hospital coding for AKI (from 16.08% below threshold, absolute increase of +5.88%, 95% CI 2.22, 7.58) but alerts did not lead to a corresponding subsequent increase in documentation of AKI in primary care (3.30% below threshold, absolute increase +0.72%, 95% CI −0.67, 1.30). There were small increases in guideline recommended post-AKI care processes including testing for proteinuria (absolute increase +4.33%, 95% CI 0.76, 7.28) and blood pressure measurement (absolute increase +3.79%, 95% CI 0.58, 6.79), which were rarely completed. There were no differences in likelihood of subsequent oral NSAID or PPI prescription, nor differences in likelihood of continued use of RAASI or diuretics among those with prescriptions prior to admission.

In the community setting, the effects of AKI alerts were similar on all outcomes to those observed in the hospital setting, with the exception of NSAID prescribing which unexpectedly increased with AKI alerts (absolute increase 3.23%, 95% CI 0.45, 6.77). While there was an increase in documentation of AKI in primary care (1.97% below threshold, absolute increase +3.27%, 95% CI 1.56, 4.25), this was again rarely completed, (table 1). The ITTs on the AKI documentation in hospital and GP are shown in Figure 2.

### Robustness checks

The effect of AKI alerts on clinical and process outcomes remained consistent across analyses using alternative bandwidth choices and regression specifications (table S8). The estimates remained similar in additional care process analyses that did not exclude patients who died during follow up (table S8). Analyses for the two negative outcome controls (new-onset cancer and statin continuation) confirmed no effect of AKI alerts on these outcomes, as expected (table S3). When the RDD analysis used a population defined by the same inclusion criteria but restricted to serum creatinine tests conducted between January 1, 2010, and January 1, 2012 (before the AKI alert algorithm was introduced in Wales), we observed no significant effects on outcomes, as expected (table S4).

### Subgroup Analyses

There were no differences in the effect of AKI alerts in a hospital setting with an interruptive vs passive design for most clinical outcomes. An interruptive alert had a larger impact on the likelihood of hospital diagnosis coding for AKI (15.55% just below threshold, absolute increase +12.66%, 95% CI 6.23, 14.94) (table 2, table S5A), with a small but statistically significant increase in subsequent documentation in primary care (5.18% just below threshold, absolute increase +3.29%, 95% CI 0.17, 4.14). The interruptive alert also led to an increase in repeat inpatient creatinine test (absolute increase +9.80%, 95% CI 1.90, 15.30), but this did not lead to a difference in effect on any kidney outcomes. Figure 3 depicts the ITTs on the health outcomes and AKI documentation by interruptive vs passive alert design. In addition, for both hospital and community settings, we detected no meaningful differential effects of AKI alerts by neighbourhood rurality, neighbourhood deprivation, sex, history of vascular disease, diabetes, or having major surgery during admission (Table. S5, Table. S6).

**Table 2.**
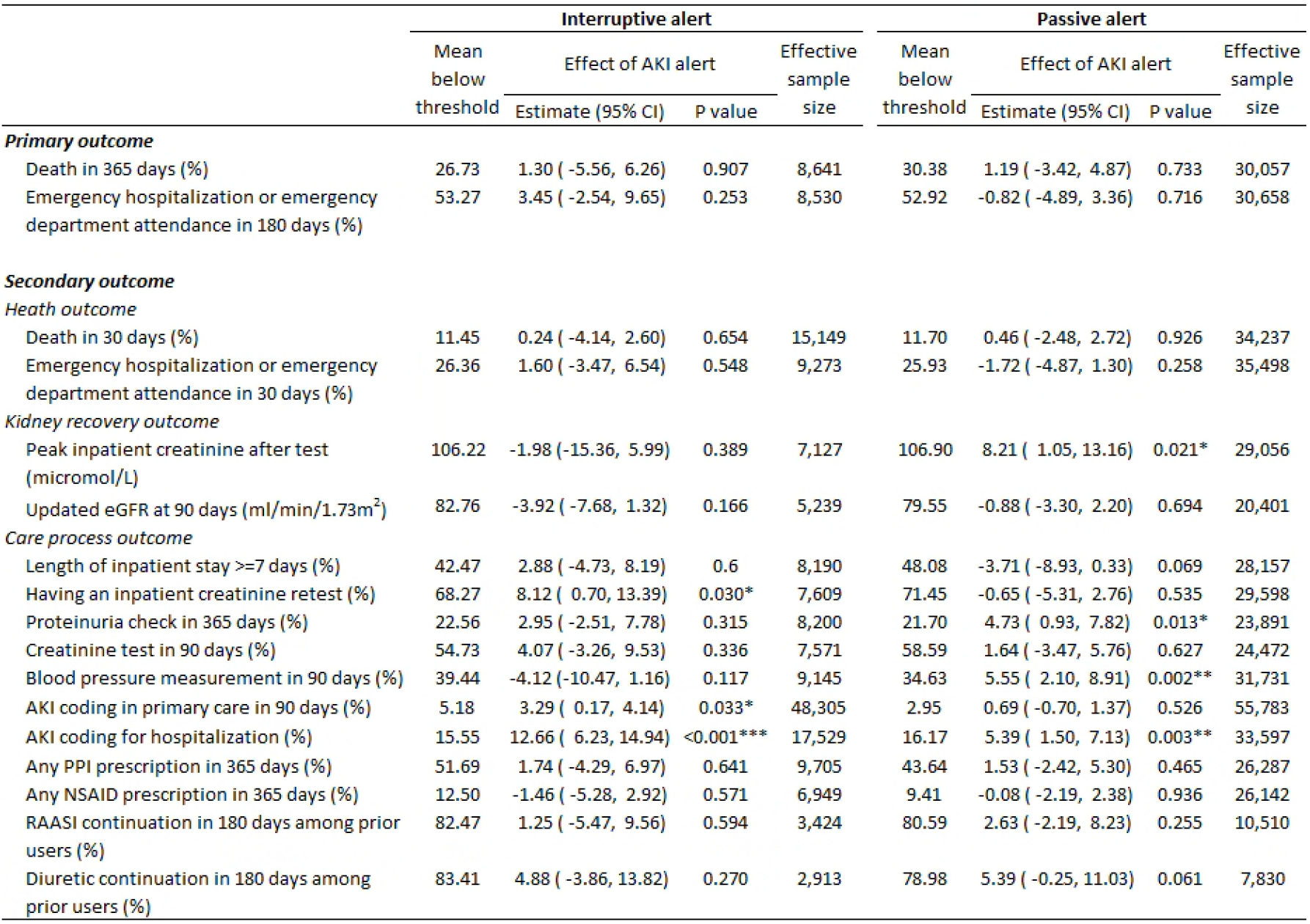
Effects of AKI alerts in emergency hospitalization setting by interruptive and passive alert approach^1,2,3^. ^1^Mean below threshold is calculated as the mean within the 5 micromol/L below the threshold. ^2^For each outcome, the MSE-optimal bandwidth is included in Panel A Table S5 in the supplement. ^3^Effective sample size is the number of observations within the MSE-optimal bandwidth. *Notes*: A positive estimate indicates the AKI alert associated with an increase in the outcome in the units of the outcome (%, unless otherwise stated)

|  | Interruptive alert |  |  |  | Passive alert |  |  |  |
| --- | --- | --- | --- | --- | --- | --- | --- | --- |
|  | Mean below threshold | Effect of AKI alert |  | Effective sample size | Mean below threshold | Effect of AKI alert |  | Effective sample size |
|  |  | Estimate (95% CI) | P value |  |  | Estimate (95% CI) | P value |  |
| **Primary outcome** |  |  |  |  |  |  |  |  |
| Death in 365 days (%) | 26.73 | 1.30 ( -5.56, 6.26) | 0.907 | 8,641 | 30.38 | 1.19 ( -3.42, 4.87) | 0.733 | 30,057 |
| Emergency hospitalization or emergency department attendance in 180 days (%) | 53.27 | 3.45 ( -2.54, 9.65) | 0.253 | 8,530 | 52.92 | -0.82 ( -4.89, 3.36) | 0.716 | 30,658 |
| **Secondary outcome** |  |  |  |  |  |  |  |  |
| *Heath outcome* |  |  |  |  |  |  |  |  |
| Death in 30 days (%) | 11.45 | 0.24 ( -4.14, 2.60) | 0.654 | 15,149 | 11.70 | 0.46 ( -2.48, 2.72) | 0.926 | 34,237 |
| Emergency hospitalization or emergency department attendance in 30 days (%) | 26.36 | 1.60 ( -3.47, 6.54) | 0.548 | 9,273 | 25.93 | -1.72 ( -4.87, 1.30) | 0.258 | 35,498 |
| *Kidney recovery outcome* |  |  |  |  |  |  |  |  |
| Peak inpatient creatinine after test (micromol/L) | 106.22 | -1.98 ( -15.36, 5.99) | 0.389 | 7,127 | 106.90 | 8.21 ( 1.05, 13.16) | 0.021* | 29,056 |
| Updated eGFR at 90 days (ml/min/1.73m<sup>2</sup>) | 82.76 | -3.92 ( -7.68, 1.32) | 0.166 | 5,239 | 79.55 | -0.88 ( -3.30, 2.20) | 0.694 | 20,401 |
| *Care process outcome* |  |  |  |  |  |  |  |  |
| Length of inpatient stay >=7 days (%) | 42.47 | 2.88 ( -4.73, 8.19) | 0.6 | 8,190 | 48.08 | -3.71 ( -8.93, 0.33) | 0.069 | 28,157 |
| Having an inpatient creatinine retest (%) | 68.27 | 8.12 ( 0.70, 13.39) | 0.030* | 7,609 | 71.45 | -0.65 ( -5.31, 2.76) | 0.535 | 29,598 |
| Proteinuria check in 365 days (%) | 22.56 | 2.95 ( -2.51, 7.78) | 0.315 | 8,200 | 21.70 | 4.73 ( 0.93, 7.82) | 0.013* | 23,891 |
| Creatinine test in 90 days (%) | 54.73 | 4.07 ( -3.26, 9.53) | 0.336 | 7,571 | 58.59 | 1.64 ( -3.47, 5.76) | 0.627 | 24,472 |
| Blood pressure measurement in 90 days (%) | 39.44 | -4.12 ( -10.47, 1.16) | 0.117 | 9,145 | 34.63 | 5.55 ( 2.10, 8.91) | 0.002** | 31,731 |
| AKI coding in primary care in 90 days (%) | 5.18 | 3.29 ( 0.17, 4.14) | 0.033* | 48,305 | 2.95 | 0.69 ( -0.70, 1.37) | 0.526 | 55,783 |
| AKI coding for hospitalization (%) | 15.55 | 12.66 ( 6.23, 14.94) | <0.001*** | 17,529 | 16.17 | 5.39 ( 1.50, 7.13) | 0.003** | 33,597 |
| Any PPI prescription in 365 days (%) | 51.69 | 1.74 ( -4.29, 6.97) | 0.641 | 9,705 | 43.64 | 1.53 ( -2.42, 5.30) | 0.465 | 26,287 |
| Any NSAID prescription in 365 days (%) | 12.50 | -1.46 ( -5.28, 2.92) | 0.571 | 6,949 | 9.41 | -0.08 ( -2.19, 2.38) | 0.936 | 26,142 |
| RAASI continuation in 180 days among prior users (%) | 82.47 | 1.25 ( -5.47, 9.56) | 0.594 | 3,424 | 80.59 | 2.63 ( -2.19, 8.23) | 0.255 | 10,510 |
| Diuretic continuation in 180 days among prior users (%) | 83.41 | 4.88 ( -3.86, 13.82) | 0.270 | 2,913 | 78.98 | 5.39 ( -0.25, 11.03) | 0.061 | 7,830 |

**Figure 3.**
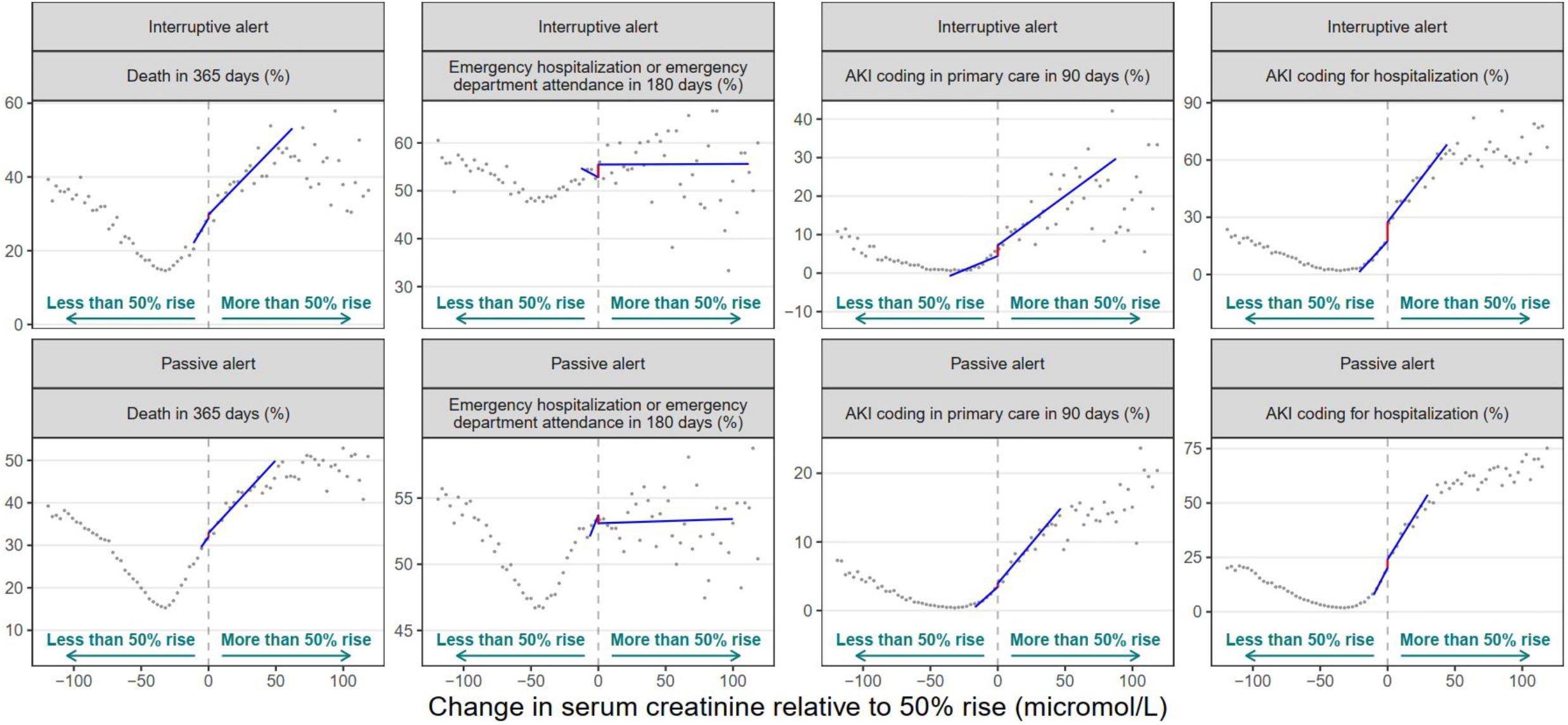
Association between the change in creatinine and outcomes by alert approach in hospital setting ^**1**,**2**^. ^1^ Grey dots show the mean value for each bin of change in creatinine. ^2^ The blue lines are fitted by our main RDD model. Red vertical lines denote the discontinuity. *Notes*: Irrespective of whether AKI alerts where delivered passively (prompt on screen) or interruptively (including phone call, ca re bundle and outreach) there was no significant difference in mortality, or readmission at the threshold for alerting. There was a larger increase in AKI coding for the hospitalisation, but this difference was modest

## Discussion

In this real-world study, we used regression discontinuity analysis as a novel approach to assess the causal impact of AKI alerts – a variably implemented electronic prompt based on a threshold trigger. At the implemented threshold in Wales (50% creatinine rise) AKI alerts had no meaningful impact on either short or long term mortality in hospital or community settings, irrespective of whether a minimal passive approach was taken, or an interruptive alert was used in combination with a care bundle. In contrast to previous real world observational studies, we also found no differences in prolonged length of hospital stay, or peak creatinine. In extended follow up after discharge, AKI alerts also did not lead to differences in admission/readmission, use of kidney protective or harmful medications, or recovery of kidney function by 90 days. At the 50% threshold, hospital and primary care coding of AKI was not frequently done, and AKI alerts made only a modest difference. Collectively the results suggest that despite a perceived clinical need to improve clinical recognition and responses (due to high mortality, subsequent admission rates, and low adherence to guideline recommended care in routine practice), nationwide alerting of AKI was largely ineffective due to a lack of meaningful impact on care processes, or outcomes.

A major strength of our approach is the simultaneous use of real-world data from a large, population-based implementation of AKI alerts in routine clinical practice outside a trial whilst retaining the ability to evaluate causal effects of an intervention through a regression discontinuity approach that mitigates the biases inherent with most other observational studies. RDD was feasible across all settings including both passive and interruptive implementation with a care bundle, and may be promising for any clinical decision support system (CDSS) evaluations that involve a trigger threshold such as a laboratory result. A RDD approach could also feasibly guide iterative fine tuning of interventions such as identify groups likely to benefit, identify intended /unintended consequences for clinical care, or demonstrate implications of a change in trigger threshold. In our example of AKI, we were able to evaluate effectiveness across different clinical settings, different approaches to alerting, a range of early and late outcomes, and with a series of robustness checks. While not applicable for the Wales implementation, this approach can be adapted to align with any changes occurring in alerts over time, in other jurisdictions such as changes in threshold trigger or provision of support.

An important consideration of RDD is that the interpretation of the findings applies specifically to the settings of the intervention as conducted. Here the nationwide AKI alert initiative relied on a 50% creatinine rise trigger threshold, which means we can only provide information on the effectiveness at this threshold. For instance, it would only be possible to address whether an alert would have been effective at a 100% creatinine rise threshold if this had been part of the original intervention, or if thresholds differed across regions within the study, or if fine tuning of the alert threshold occurred over time during the observed period under study. Similarly, while we report serious under-coding of AKI with only small improvements among those who received alerts, we cannot directly ascertain if this represents a lack of documentation despite clinical recognition and action, or a lack of perceived importance of the alert itself leading to inaction. However, by evaluating a range of care process and documentation measures, we noted a consistent lack of change in clinical action across a range of processes which implies the latter. In addition, the intent of AKI alerts was to drive improvements in recognition and early care.

Therefore, changes in post-discharge care after AKI might be regarded as beyond remit, although we note that documentation and post-AKI care were a component of clinical guidelines and emphasised good practice as part of the Think Kidneys primary care resources in April 2016. Finally, as with a clinical trial, a lack of effect with RDD design may represent a type II error: a failure to detect an effect that is present, such as due to insufficient sample size or event rate. As reported in our pre-registered protocol, we had limited power to evaluate NSAID and diuretic use after AKI although these are important care processes. Nonetheless, our findings suggest if any effects existed on these processes of care, they are likely to be small.

This study adds insight to the conflicting findings from trials and observational studies of AKI alerts, with findings that cover hospital and the community settings and both short and long-term horizons. Consistent with several randomised trials^25, 26^, and contrary to several observation studies^16–19, 22, 23^, our study found a lack of effect on mortality, and additionally found no meaningful effect from alerts (as implemented) on kidney recovery, or emergency hospital attendances, admissions, and readmissions. Our findings favour the generalisability of trial findings to real-world settings over the possibility that real-world “before and after studies”identified small effects that went undetected in previous trials. We also assessed previous trial findings of reductions in risk of prolonged hospitalisation^24^, potential for harm among patients undergoing surgery and potential for harm in non-teaching hospitals^26^ – for which we found no evidence of beneficial or harmful effect.

This study also highlights a promising role for RDD for the evaluation of future CDSS-based interventions. A recent systematic review of computerised decision support found that uptake was rarely reported and generally low when evaluated, which precludes learning for future interventions^8^. Our RDD approach enabled us to evaluate both the benefit and response in the real-world^8^. This approach could be repeated with iterative fine-tuning. Future iteration of the current approach in Wales, for instance, could involve (1) a change in threshold trigger or patient subset to cover those with greater need, (2) targeted recommendations based on medications to minimise nephrotoxicity, or to avoid harm from stopping or failing to restart medications, (3) greater intensity and resource for those most likely to benefit from an intervention, or (4) additional prompts to integrate assessment of kidney recovery in the community^21, 39-42^. Such tailored approaches may be especially promising given the expanding therapeutic options available for people with new onset albuminuric CKD or unrecovered AKI^43, 44^.

In conclusion, in a real-world regression discontinuity evaluation of a nationwide implementation of AKI alerts, we found no evidence for benefit of widespread alerts in hospital or community settings, with either interruptive or passive approaches, or in any clinical subgroups. At the implemented 50% creatinine threshold, even an interruptive alert with a care bundle had little impact on subsequent documentation of an AKI diagnosis in hospital or primary care. Thus, the NHS Wales initiative as implemented appears to have been insufficient to improve the care and outcomes for people with AKI. The poor outcomes and shortfalls within existing AKI care despite alerting strategies point towards a continued need for improvement and the regression discontinuity approach we have demonstrated shows promise to facilitate future real-world causal evaluations.

## Data Availability

All data produced in the present study are not publicly available. Data access would require approval by SAIL's Information Governance Review Panel.

## Acknowledgments

This study makes use of anonymised data held in the Secure Anonymised Information Linkage (SAIL) Databank. We would like to acknowledge all the data providers who make anonymised data available for research. The responsibility for the interpretation of the data supplied by SAIL is that of the authors alone. SAIL bears no responsibility for the further analysis or interpretation of their data, over and above that published by SAIL.

## Contributors

Contributors: MX and SS are joint corresponding authors. PG obtained funding. MX, PG and SS conceived the idea. MX and SS designed the study with critical input from PG, TS and MJ. MX analysed the data. MX and SS drafted the first manuscript with assistance of ZT and TB for the interpretation. MX, PG, TB, MJ, TS, ZT, and SS critically revised the manuscript for important intellectual content. All the authors approved the final manuscript. MX and SS are the guarantors. The corresponding authors (MX and SS) attest that all listed authors meet the authorship criteria and that no others that meet the criteria have been omitted.

## Data Sharing

Data access would require approval by SAIL’s Information Governance Review Panel. Information on how researchers may make requests to obtain similar datasets from the health research dataset custodians may be provided upon request.

