## Supplementary material for "Effects of nationwide alerting for acute kidney injury on healthcare and patient outcomes: population based, regression discontinuity analysis"

### Supplement

This supplement contains:

- **Additional Methods**
- **Figure S1 to S4**
- **Tables S1 to S8**

#### Additional methods

##### Running variable derivation:

For this analysis, we defined a centered running variable (RV) based on the AKI algorithm as implemented in Wales: for an index serum creatinine measurement on a given date (C), the baseline value (B) was determined as the minimum serum creatinine level recorded in the preceding seven days. If no tests were conducted in this timeframe, the baseline value was calculated as the median value from the preceding 8–365 days. The RV was then calculated as $RV=C-(1.5\times B)$. Thus, the RV represented the change in creatinine level relative to the 50% rise threshold. Following the AKI alert algorithm, an alert would be triggered when RV >= 0 on the date of the index test.

##### Local linear regression:

To quantify the discontinuity, we modelled the outcomes as a linear function of the centered RV, separately for observations on either side of the threshold. The discontinuity was defined as the difference between the two regression lines at the threshold. To minimize bias from misspecifying the functional form, we restricted the analysis to observations within a narrow bandwidth around the threshold. While a narrow bandwidth reduces bias, it increases the variance of the estimate. Following the standard practice, we selected a bandwidth optimized to balance this bias-variance trade-off. Asymmetry in the running variable's distribution was accounted for by allowing different bandwidths on either side of the threshold. We further applied a triangular kernel, assigning linearly greater weight to observations closer to the threshold.

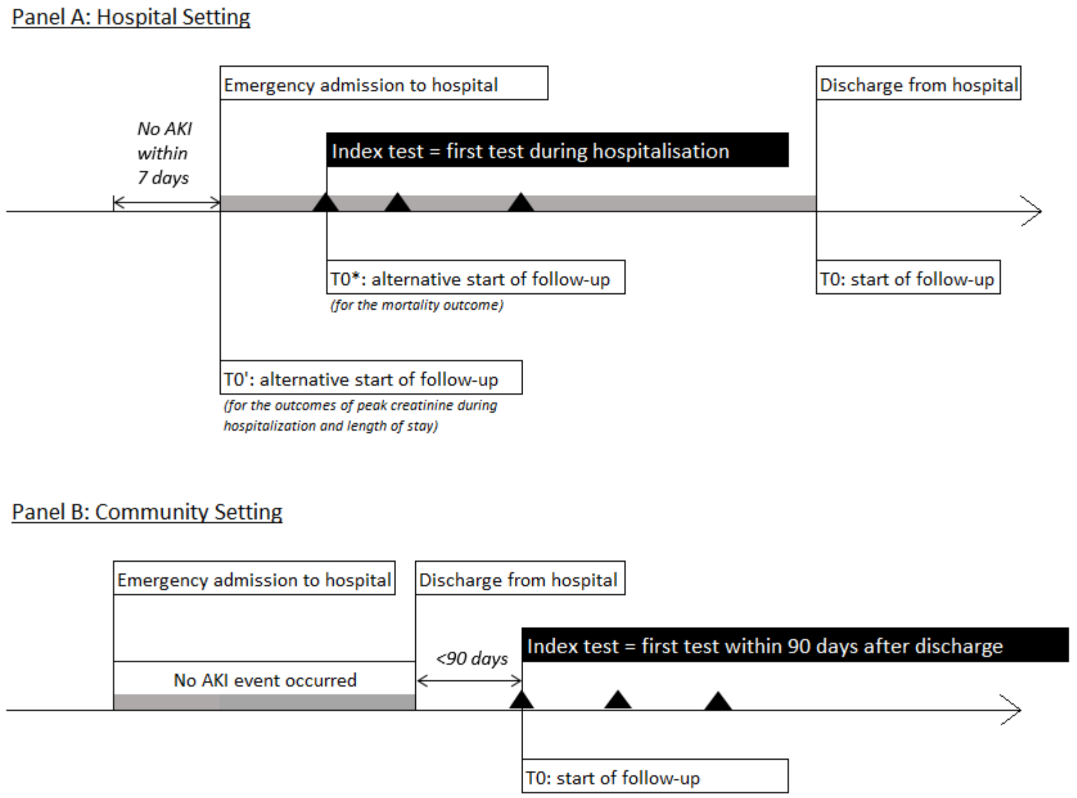

**Fig S1. Illustration of index test selection and timing of follow-up[1,2]**

[1] The triangles indicate the serum creatinine tests taken.

[2] The section in grey indicates the duration of an emergency inpatient stay.

| STOP AKI | Response | |
|---|---|---|
| Sepsis | Deliver Sepsis Six (if appropriate) | ☐ |
| Toxins | Review Medications | ☐ |
| | Stop / Avoid potential nephrotoxins ✱ | ☐ |
| Optimise BP | Assess volume status: give fluids (if appropriate) | ☐ |
| | Start a fluid balance chart | ☐ |
| | Review BP lowering medication ✱ | ☐ |
| Prevent harm | Treat complications | |
| | Monitor renal function to assess response | |
| | Record cause of AKI: | |
| ✱ Review daily - consider restarting within clinical context (including at discharge from hospital) | | |
| Urine dipstick result: | | ☐ |
| USS KUB urgent if: | • No identified cause of AKI<br>• Possibility of renal tract obstruction<br>• Deteriorating renal function | |
| Date: | Time: | |
| Name: | Grade: | Sign: |

**Fig S2. Care bundle that was completed and recorded for interruptive alerts.**

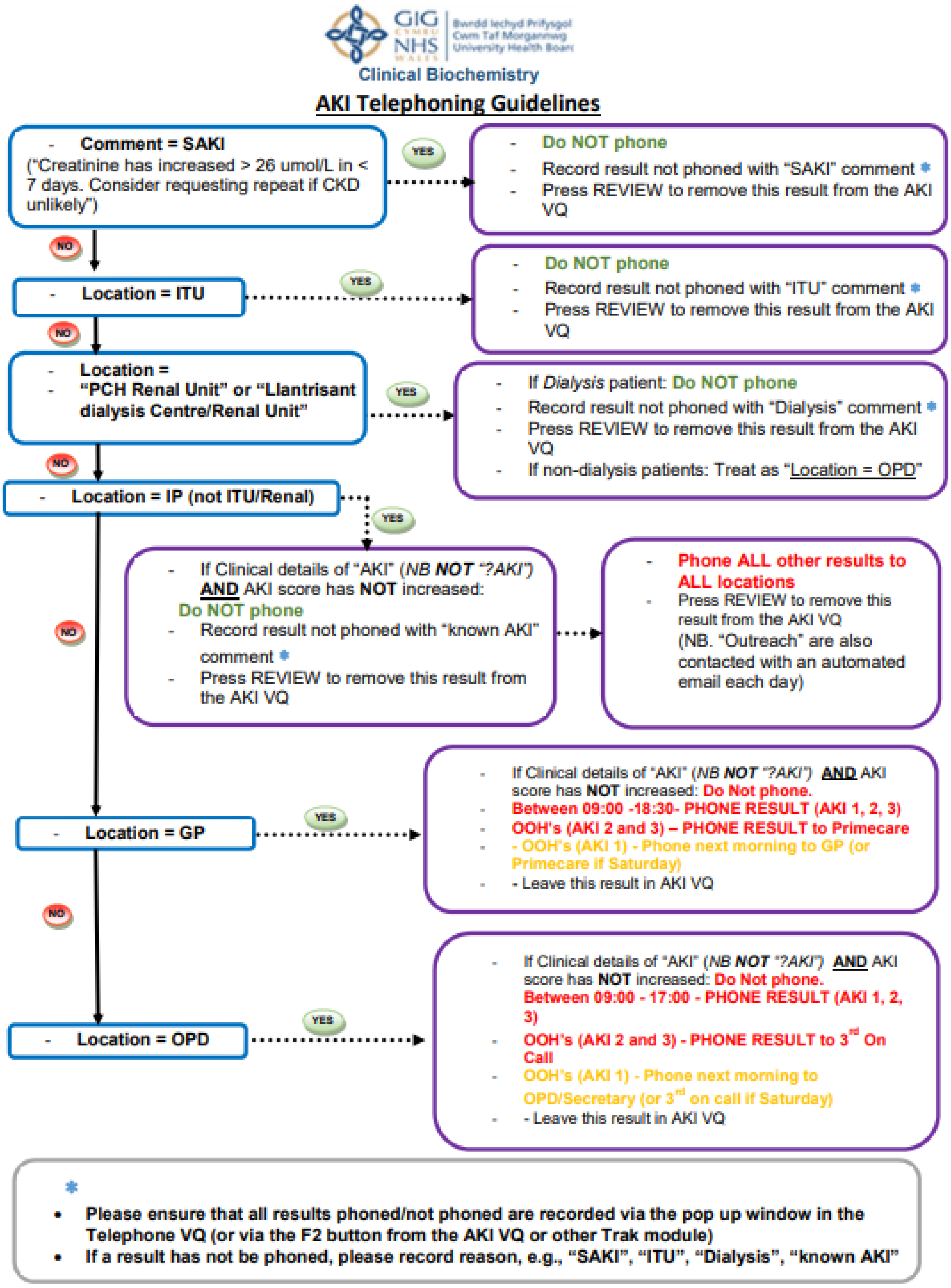

**Fig S3. Algorithm used for the interruptive telephoning of AKI alerts.**

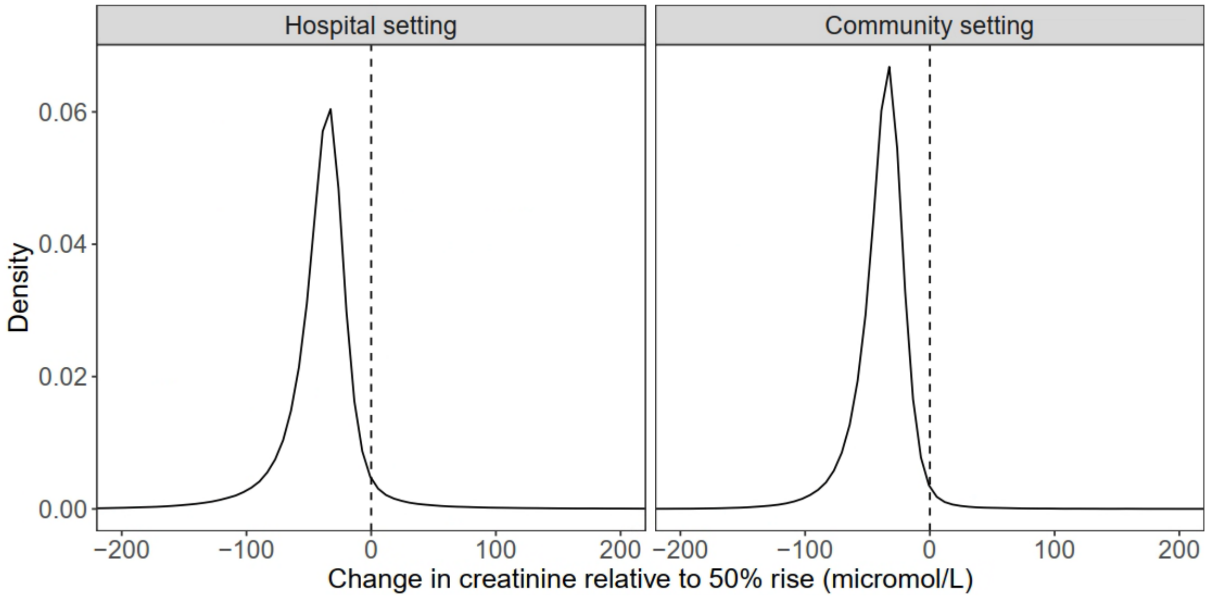

**Fig S4. Distribution of the centred running variable for the hospital and community setting.**

| | Hospital setting | | | | Community setting | | | |
|---|---|---|---|---|---|---|---|---|
| | N | Mean: Full sample | Mean: Sample near threshold: below | Mean: Sample near threshold: above | N | Mean: Full sample | Mean: Sample near threshold: below | Mean: Sample near threshold: above |
| ***Baseline variables*** | | | | | | | | |
| Age (year) | 861,494 | 64.64 | 66.94 | 68.28 | 354,505 | 64.25 | 64.55 | 66.06 |
| Female (%) | 861,491 | 54.47 | 60.68 | 57.00 | 354,505 | 54.41 | 63.60 | 60.26 |
| Rural area (%) | 861,494 | 32.61 | 33.64 | 33.46 | 354,505 | 33.30 | 34.61 | 34.22 |
| WIMD decile (1 = most deprived) | 816,252 | 5.05 | 4.97 | 4.98 | 335,528 | 5.13 | 5.07 | 5.05 |
| Baseline eGFR (ml/min/1.73m$^2$) | 861,494 | 78.20 | 85.78 | 81.66 | 354,505 | 81.15 | 92.78 | 89.51 |
| Had diabetes at baseline (%) | 861,494 | 23.60 | 27.63 | 30.50 | 354,505 | 24.70 | 24.78 | 27.90 |
| Had CKD3+ at baseline (%) | 861,494 | 29.26 | 23.28 | 27.60 | 354,505 | 24.69 | 17.55 | 21.07 |
| Had heart failure at baseline (%) | 861,494 | 16.67 | 19.84 | 21.66 | 354,505 | 16.26 | 18.28 | 20.27 |
| Had cerebrovascular disease in the past 180 days (% | 861,494 | 2.82 | 2.39 | 2.60 | 354,505 | 3.88 | 2.75 | 3.36 |
| Had peripheral artery disease in the past 180 days (9 | 861,494 | 3.15 | 3.89 | 3.81 | 354,505 | 2.86 | 3.75 | 4.16 |
| Had coronary heart disease in the past 180 days (%) | 861,494 | 19.06 | 19.34 | 19.98 | 354,505 | 19.84 | 17.55 | 17.86 |
| Had stroke in the past 180 days (%) | 861,494 | 4.12 | 3.84 | 4.00 | 354,505 | 3.88 | 2.82 | 3.61 |
| RAASI use in the past 180 days (%) | 861,494 | 28.04 | 30.47 | 33.07 | 354,505 | 31.23 | 28.43 | 30.21 |
| Diuretic use in the past 180 days (%) | 861,494 | 17.01 | 20.93 | 23.04 | 354,505 | 18.47 | 23.29 | 23.83 |
| NSAID use in the past 180 days (%) | 861,494 | 8.99 | 8.35 | 8.01 | 354,505 | 10.07 | 9.06 | 8.88 |
| PPI use in the past 180 days (%) | 861,494 | 38.60 | 40.80 | 40.78 | 354,505 | 42.84 | 43.76 | 43.75 |
| Statin use in the past 180 days (%) | 861,494 | 31.45 | 32.68 | 34.69 | 354,505 | 34.31 | 29.56 | 32.36 |
| Interruptive alert setting (%) | 861,494 | 14.06 | 15.55 | 15.54 | | | | |
| With history of vascular disease (%) | 861,494 | 31.27 | 33.80 | 35.73 | 354,505 | 32.48 | 31.25 | 33.82 |
| With major surgery during admission (%) | 861,494 | 0.78 | 0.92 | 0.94 | | | | |
| ***Exposure variable*** | | | | | | | | |
| AKI alert (%) | 861,494 | 5.75 | 16.52 | 92.70 | 354,505 | 2.01 | 5.84 | 91.22 |
| ***Outcome variables*** | | | | | | | | |
| Death in 365 days (%) | 861,494 | 21.14 | 29.81 | 32.54 | 354,505 | 15.79 | 25.32 | 28.65 |
| Emergency hospitalization or emergency department attendance in 180 days (%) | 808,890 | 49.26 | 52.98 | 53.56 | 354,505 | 51.06 | 61.18 | 66.13 |
| Death in 30 days (%) | 861,494 | 6.33 | 11.66 | 13.83 | 354,505 | 2.60 | 6.64 | 8.63 |
| Emergency hospitalization or emergency department attendance in 30 days (%) | 808,890 | 23.48 | 26.00 | 26.13 | 354,505 | 26.68 | 40.15 | 45.86 |
| Having an inpatient creatinine retest (%) | 861,494 | 54.88 | 70.95 | 77.31 | | | | |
| Peak inpatient creatinine after test (micromol/L) | 472,763 | 104.76 | 106.80 | 116.23 | | | | |
| Updated eGFR at 90 days (ml/min/1.73m2) | 589,545 | 79.08 | 80.04 | 77.10 | 212,213 | 78.77 | 81.51 | 78.21 |
| Length of inpatient stay >=7 days (%) | 843,778 | 37.00 | 47.21 | 50.28 | | | | |
| Any PPI prescription in 365 days (%) | 675,317 | 43.16 | 44.95 | 45.37 | 298,524 | 45.63 | 47.80 | 46.77 |
| Any NSAID prescription in 365 days (%) | 675,317 | 11.08 | 9.91 | 9.34 | 298,524 | 10.97 | 8.97 | 10.06 |
| RAASI continuation in 180 days among prior users | 198,243 | 84.82 | 80.93 | 80.45 | 99,965 | 88.36 | 85.90 | 83.55 |
| Diuretic continuation in 180 days among prior users | 105,081 | 82.79 | 79.72 | 82.37 | 54,709 | 84.39 | 82.99 | 82.84 |
| Statin continuation in 180 days among prior users | 220,755 | 91.82 | 90.03 | 92.07 | 108,962 | 93.07 | 92.19 | 89.21 |
| Having cancer in 365 days (%) | 808,890 | 3.64 | 3.55 | 3.70 | 354,505 | 3.02 | 2.69 | 3.31 |
| Proteinuria check in 365 days (%) | 675,317 | 20.00 | 21.84 | 24.34 | 298,524 | 21.94 | 21.50 | 24.61 |
| Creatinine test in 90 days (%) | 752,412 | 52.78 | 57.98 | 59.08 | 332,115 | 55.19 | 66.29 | 68.19 |
| Blood pressure measurement in 90 days (%) | 752,412 | 34.02 | 35.38 | 37.65 | 332,115 | 36.43 | 36.56 | 36.32 |
| AKI coding in primary care in 90 days (%) | 760,553 | 1.54 | 3.30 | 4.67 | 332,115 | 0.59 | 1.97 | 5.45 |
| AKI coding for hospitalization (%) | 808,890 | 6.18 | 16.08 | 26.48 | | | | |

#### Table S1. sample characteristics[1]

[1]”Sample near threshold” is defined as observations with a change in serum creatinine relative to its 50% increase threshold within [-5, 0) and [0, 5) micromol/L for “below” and “above” the threshold, respectively.

[2]N is the sample size. There are 861,494 observations included in the hospital setting, among with, 52,604 observations are excluded for emergency hospitalization or emergency department attendance and AKI coding for hospitalization outcomes because they did not survive until discharge; 388,731 are excluded for peak inpatient creatinine outcome because they did not receive another creatinine test during the hospitalization; 271,949 are excluded for having updated eGFR outcome because they did not test for eGFR or survive within 90 days; 17,716 are excluded for length of stay outcome because they did not survive for at least 7 days; 186,177 are excluded for PPI and NSAID prescription, and proteinuria check outcomes because they did not survive for at least 365 days; 663,251, 756,413 and 640,739 are excluded for RAASI, diuretic and statin continuation outcomes, respectively, because they were not prescribed with the respective medication within 180 days before test or did not survive for at least 180 days after discharge; 109,082

are excluded for creatinine test and blood pressure measurement because they did not survive for at least 90 days after discharge; 100,941 are excluded for AKI coding in primary care because they did not survive until discharge or for at least 90 days after test.

[3]There are 354,505 observations included in the community setting, among with, 142,292 are excluded for having updated eGFR outcome because they did not test for eGFR or survive within 90 days; 55,981 are excluded for PPI and NSAID prescription, and proteinuria check outcomes because they did not survive for at least 365 days; 254,540, 299,796 and 245,543 are excluded for RAASI, diuretic and statin continuation outcomes, respectively, because they were not prescribed with the respective medication within 180 days before test or did not survive for at least 180 days after test; 22,390 are excluded for creatinine test, blood pressure measurement and AKI coding in primary care because they did not survive for at least 90 days after test.

| | Hospital setting | | | | | | Community setting | | | | | |
|---|---|---|---|---|---|---|---|---|---|---|---|---|
| | Mean below threshold | Difference at the cutoff | | Bandwidth (micromol/L) | | Effective sample size | Mean below threshold | Difference at the cutoff | | Bandwidth (micromol/L) | | Effective sample size |
| | | Estimate (95% CI) | P value | below | above | | | Estimate (95% CI) | P value | below | above | |
| ***Exposure variable*** | | | | | | | | | | | | |
| AKI alert (%) | 16.52 | 71.52 (68.63, 73.70) | <0.001*** | 5 | 38 | 30,796 | 5.84 | 82.88 (79.78, 85.00) | <0.001*** | 7 | 38 | 8,872 |
| ***Baseline variable*** | | | | | | | | | | | | |
| Had diabetes at baseline (%) | 27.63 | 1.50 (-0.40, 2.83) | 0.140 | 14 | 68 | 66,391 | 24.78 | 1.50 (-2.18, 4.06) | 0.554 | 12 | 55 | 17,572 |
| Had CKD3+ at baseline (%) | 23.28 | 2.02 (-0.48, 4.00) | 0.123 | 8 | 36 | 37,488 | 17.55 | -0.98 (-5.90, 2.38) | 0.405 | 6 | 36 | 8,792 |
| Had heart failure at baseline (%) | 19.84 | 0.60 (-1.32, 1.86) | 0.743 | 11 | 55 | 55,230 | 18.28 | -0.41 (-3.96, 1.92) | 0.497 | 11 | 46 | 15,485 |
| Had cerebrovascular disease in the past 180 days (%) | 2.39 | 0.10 (-0.38, 0.51) | 0.779 | 18 | 167 | 104,929 | 2.75 | 0.57 (-0.42, 1.42) | 0.287 | 23 | 87 | 54,476 |
| Had peripheral artery disease in the past 180 days (%) | 3.89 | -0.20 (-0.94, 0.33) | 0.351 | 17 | 78 | 85,473 | 3.75 | 0.30 (-1.08, 1.31) | 0.852 | 16 | 72 | 24,875 |
| Had coronary heart disease in the past 180 days (%) | 19.34 | 0.68 (-0.98, 2.06) | 0.486 | 12 | 100 | 64,211 | 17.55 | -0.90 (-4.35, 1.48) | 0.333 | 11 | 46 | 15,518 |
| Had stroke in the past 180 days (%) | 3.84 | 0.29 (-0.34, 0.92) | 0.369 | 16 | 111 | 81,136 | 2.82 | 0.80 (-0.25, 1.69) | 0.145 | 22 | 81 | 54,442 |
| Age (year) | 66.94 | 0.20 (-0.78, 0.84) | 0.937 | 11 | 37 | 51,775 | 64.55 | 0.19 (-1.52, 1.29) | 0.873 | 11 | 44 | 15,429 |
| Female (%) | 60.68 | -2.34 (-4.54, 0.55) | 0.125 | 7 | 62 | 42,460 | 63.60 | -3.35 (-7.26, 1.54) | 0.203 | 8 | 67 | 10,574 |
| RAASI use in the past 180 days (%) | 30.47 | 1.28 (-1.01, 2.95) | 0.337 | 10 | 65 | 52,650 | 28.43 | -0.16 (-4.32, 2.86) | 0.690 | 10 | 53 | 14,008 |
| Diuretic use in the past 180 days (%) | 20.93 | 0.38 (-1.54, 1.62) | 0.960 | 12 | 54 | 59,464 | 23.29 | -2.50 (-6.39, 0.01) | 0.051 | 11 | 44 | 15,457 |
| NSAID use in the past 180 days (%) | 8.35 | -0.62 (-1.59, 0.28) | 0.168 | 14 | 137 | 76,389 | 9.06 | 0.21 (-1.47, 1.98) | 0.773 | 17 | 90 | 27,947 |
| PPI use in the past 180 days (%) | 40.80 | 0.01 (-1.76, 1.32) | 0.780 | 17 | 96 | 94,002 | 43.76 | 0.68 (-2.43, 3.29) | 0.770 | 18 | 97 | 31,293 |
| Statin use in the past 180 days (%) | 32.68 | 1.71 (-0.38, 3.44) | 0.117 | 11 | 81 | 58,412 | 29.56 | 1.04 (-2.95, 4.00) | 0.767 | 11 | 60 | 15,831 |
| Baseline eGFR (ml/min/1.73m$^2$) | 85.78 | -1.26 (-2.40, 0.55) | 0.217 | 7 | 28 | 35,004 | 92.78 | 1.08 (-1.11, 4.48) | 0.237 | 6 | 31 | 8,627 |
| Rural area (%) | 33.64 | -0.09 (-1.63, 1.77) | 0.936 | 12 | 156 | 66,575 | 34.61 | -1.19 (-4.11, 2.22) | 0.560 | 13 | 86 | 20,056 |
| WIMD decile (1 = most deprived) | 4.97 | 0.03 (-0.08, 0.11) | 0.721 | 15 | 107 | 76,416 | 5.07 | 0.03 (-0.12, 0.20) | 0.616 | 22 | 80 | 51,291 |

#### Table S2. Balance tests for baseline variables[1,2,3]

[1]Mean below threshold is calculated as the mean within the 5 micromol/L below the threshold.

[2] Effective sample size is the number of observations within the MSE-optimal bandwidth.

[3] Abbreviations: AKI = acute kidney injury; CKD3+ = chronic kidney disease of stage 3 or higher; WIMD decile = Welsh Index of Multiple Deprivation decile (1 = most deprived).

| | Hospital setting | | | | | | Community setting | | | | | |
|---|---|---|---|---|---|---|---|---|---|---|---|---|
| | Mean below threshold | Effect of AKI alert | | Bandwidth (micromol/L) | | Effective sample size | Mean below threshold | Effect of AKI alert | | Bandwidth (micromol/L) | | Effective sample size |
| | | Estimate (95% CI) | P value | below | above | | | Estimate (95% CI) | P value | below | above | |
| ***Negative outcome control*** | | | | | | | | | | | | |
| Statin continuation in 180 days among prior users (%) | 90.03 | 1.37 (-1.52, 4.53) | 0.330 | 11 | 77 | 14,390 | 92.19 | -3.44 (-7.70, 0.77) | 0.108 | 14 | 69 | 5,437 |
| Having cancer in 365 days (%) | 3.55 | -0.09 (-1.24, 0.86) | 0.720 | 11 | 98 | 47,454 | 2.69 | 0.51 (-0.84, 1.76) | 0.489 | 13 | 101 | 20,184 |

#### Table S3. Negative outcome controls[1,2,3]

[1]Mean below threshold is calculated as the mean within the 5 micromol/L below the threshold.

[2] Effective sample size is the number of observations within the MSE-optimal bandwidth.

[3] Abbreviations: AKI = acute kidney injury

| | Hospital setting | | | | | | Community setting | | | | | |
|---|---|---|---|---|---|---|---|---|---|---|---|---|
| | Mean below threshold | Effect of AKI alert | | Bandwidth | | Effective sample size | Mean below threshold | Effect of AKI alert | | Bandwidth | | Effective sample size |
| | | Estimate (95% CI) | P value | below | above | | | Estimate (95% CI) | P value | below | above | |
| ***Primary outcome*** | | | | | | | | | | | | |
| Death in 365 days (%) | 31.60 | 1.57 (-0.93, 3.16) | 0.284 | 17 | 44 | 49,077 | 24.51 | 1.62 (-3.20, 4.96) | 0.672 | 13 | 58 | 10,440 |
| Emergency hospitalization or emergency department attendance in 180 days (%) | 50.43 | 1.78 (-0.40, 4.25) | 0.105 | 13 | 149 | 37,354 | 54.94 | 2.35 (-2.49, 5.53) | 0.458 | 17 | 62 | 16,057 |
| ***Secondary outcome*** | | | | | | | | | | | | |
| *Heath outcome* | | | | | | | | | | | | |
| Death in 30 days (%) | 12.95 | 1.83 (-0.12, 3.16) | 0.069 | 14 | 45 | 41,874 | 5.87 | 1.76 (-0.93, 3.67) | 0.243 | 14 | 67 | 11,730 |
| Emergency hospitalization or emergency department attendance in 30 days (%) | 24.50 | -0.58 (-2.14, 0.72) | 0.33 | 30 | 133 | 138,110 | 33.19 | 3.97 (-0.95, 7.40) | 0.131 | 15 | 68 | 11,740 |
| *Kidney recovery outcome* | | | | | | | | | | | | |
| Peak inpatient creatinine after test (micromol/L) | 111.40 | 3.21 (-1.91, 7.19) | 0.255 | 12 | 62 | 26,424 | | | | | | |
| Updated eGFR at 90 days (ml/min/1.73m$^2$) | 77.64 | -1.24 (-2.97, 0.86) | 0.282 | 10 | 45 | 20,663 | 77.08 | 3.15 (-0.52, 8.67) | 0.082 | 7 | 43 | 3,417 |
| *Care process outcome* | | | | | | | | | | | | |
| Length of inpatient stay >=7 days (%) | 51.98 | 1.45 (-1.06, 3.56) | 0.290 | 15 | 47 | 42,929 | | | | | | |
| Having an inpatient creatinine retest (%) | 69.46 | 1.29 (-0.96, 3.11) | 0.301 | 16 | 39 | 44,595 | | | | | | |
| Proteinuria check in 365 days (%) | 25.58 | 0.32 (-2.66, 2.44) | 0.933 | 12 | 74 | 24,784 | 24.68 | 0.64 (-5.13, 4.97) | 0.975 | 11 | 75 | 5,542 |
| Creatinine test in 90 days (%) | 50.89 | 0.41 (-2.52, 2.40) | 0.962 | 14 | 73 | 33,627 | 57.45 | 1.78 (-3.50, 5.48) | 0.666 | 15 | 69 | 11,434 |
| Blood pressure measurement in 90 days (%) | 36.33 | -0.43 (-3.21, 1.60) | 0.513 | 14 | 92 | 34,331 | 37.23 | 0.98 (-4.26, 4.80) | 0.907 | 15 | 70 | 10,231 |
| AKI coding in primary care in 90 days (%) | 0.37 | 0.03 (-0.30, 0.28) | 0.937 | 23 | 69 | 74,663 | 0.34 | 0.21 (-0.25, 0.75) | 0.330 | 32 | 56 | 63,817 |
| AKI coding for hospitalization (%) | 4.72 | 1.30 (-0.31, 2.41) | 0.130 | 10 | 53 | 26,937 | | | | | | |
| Any PPI prescription in 365 days (%) | 43.43 | 2.46 (-0.01, 4.62) | 0.051 | 20 | 102 | 45,801 | 45.00 | 4.25 (-0.48, 8.15) | 0.082 | 20 | 107 | 18,011 |
| Any NSAID prescription in 365 days (%) | 13.24 | -0.24 (-1.79, 1.63) | 0.927 | 17 | 88 | 34,906 | 12.86 | 0.73 (-2.14, 4.08) | 0.541 | 17 | 114 | 12,630 |
| RAASI continuation in 180 days among prior users (%) | 83.37 | -0.15 (-2.44, 3.21) | 0.790 | 20 | 85 | 15,567 | 84.69 | 3.78 (-0.94, 9.59) | 0.108 | 19 | 84 | 4,952 |
| Diuretic continuation in 180 days among prior users (%) | 84.65 | 1.87 (-1.13, 4.60) | 0.236 | 34 | 101 | 23,177 | 87.70 | 1.49 (-4.38, 6.18) | 0.738 | 25 | 75 | 5,519 |

#### Table S4. Falsification test (Negative exposure control)[1,2,3]

[1]Mean below threshold is calculated as the mean within the 5 micromol/L below the threshold.

[2] Effective sample size is the number of observations within the MSE-optimal bandwidth.

[3] Abbreviations: AKI = acute kidney injury

**A**

| | Interruptive alert | | | | | | Passive alert | | | | | |
|---|---|---|---|---|---|---|---|---|---|---|---|---|
| | Mean below threshold | Effect of AKI alert | | Bandwidth (micromol/L) | | Effective sample size | Mean below threshold | Effect of AKI alert | | Bandwidth (micromol/L) | | Effective sample size |
| | | Estimate (95% CI) | P value | below | above | | | Estimate (95% CI) | P value | below | above | |
| ***Primary outcome*** | | | | | | | | | | | | |
| Death in 365 days (%) | 26.73 | 1.30 ( -5.56, 6.26) | 0.907 | 11 | 62 | 8,641 | 30.38 | 1.19 ( -3.42, 4.87) | 0.733 | 6 | 50 | 30,057 |
| Emergency hospitalization or emergency department attendance in 180 days (%) | 53.27 | 3.45 ( -2.54, 9.65) | 0.253 | 13 | 112 | 8,530 | 52.92 | -0.82 ( -4.89, 3.36) | 0.716 | 6 | 100 | 30,658 |
| ***Secondary outcome*** | | | | | | | | | | | | |
| *Heath outcome* | | | | | | | | | | | | |
| Death in 30 days (%) | 11.45 | 0.24 ( -4.14, 2.60) | 0.654 | 19 | 59 | 15,149 | 11.70 | 0.46 ( -2.48, 2.72) | 0.926 | 7 | 48 | 34,237 |
| Emergency hospitalization or emergency department attendance in 30 days (%) | 26.36 | 1.60 ( -3.47, 6.54) | 0.548 | 14 | 121 | 9,273 | 25.93 | -1.72 ( -4.87, 1.30) | 0.258 | 8 | 114 | 35,498 |
| *Kidney recovery outcome* | | | | | | | | | | | | |
| Peak inpatient creatinine after test (micromol/L) | 106.22 | -1.98 (-15.36, 5.99) | 0.389 | 13 | 83 | 7,127 | 106.90 | 8.21 ( 1.05, 13.16) | 0.021* | 7 | 86 | 29,056 |
| Updated eGFR at 90 days (ml/min/1.73m$^2$) | 82.76 | -3.92 ( -7.68, 1.32) | 0.166 | 10 | 61 | 5,239 | 79.55 | -0.88 ( -3.30, 2.20) | 0.694 | 6 | 43 | 20,401 |
| *Care process outcome* | | | | | | | | | | | | |
| Length of inpatient stay >=7 days (%) | 42.47 | 2.88 ( -4.73, 8.19) | 0.6 | 12 | 65 | 8,190 | 48.08 | -3.71 ( -8.93, 0.33) | 0.069 | 5 | 51 | 28,157 |
| Having an inpatient creatinine retest (%) | 68.27 | 8.12 ( 0.70, 13.39) | 0.030* | 9 | 71 | 7,609 | 71.45 | -0.65 ( -5.31, 2.76) | 0.535 | 5 | 46 | 29,598 |
| Proteinuria check in 365 days (%) | 22.56 | 2.95 ( -2.51, 7.78) | 0.315 | 15 | 134 | 8,200 | 21.70 | 4.73 ( 0.93, 7.82) | 0.013* | 8 | 81 | 23,891 |
| Creatinine test in 90 days (%) | 54.73 | 4.07 ( -3.26, 9.53) | 0.336 | 13 | 88 | 7,571 | 58.59 | 1.64 ( -3.47, 5.76) | 0.627 | 6 | 79 | 24,472 |
| Blood pressure measurement in 90 days (%) | 39.44 | -4.12 (-10.47, 1.16) | 0.117 | 14 | 115 | 9,145 | 34.63 | 5.55 ( 2.10, 8.91) | 0.002** | 9 | 129 | 31,731 |
| AKI coding in primary care in 90 days (%) | 5.18 | 3.29 ( 0.17, 4.14) | 0.033* | 36 | 88 | 48,305 | 2.95 | 0.69 ( -0.70, 1.37) | 0.526 | 17 | 47 | 55,783 |
| AKI coding for hospitalization (%) | 15.55 | 12.66 ( 6.23, 14.94) | <0.001*** | 21 | 44 | 17,529 | 16.17 | 5.39 ( 1.50, 7.13) | 0.003** | 10 | 30 | 33,597 |
| Any PPI prescription in 365 days (%) | 51.69 | 1.74 ( -4.29, 6.97) | 0.641 | 17 | 128 | 9,705 | 43.64 | 1.53 ( -2.42, 5.30) | 0.465 | 8 | 100 | 26,287 |
| Any NSAID prescription in 365 days (%) | 12.50 | -1.46 ( -5.28, 2.92) | 0.571 | 13 | 147 | 6,949 | 9.41 | -0.08 ( -2.19, 2.38) | 0.936 | 8 | 94 | 26,142 |
| RAASI continuation in 180 days among prior users (%) | 82.47 | 1.25 ( -5.47, 9.56) | 0.594 | 16 | 83 | 3,424 | 80.59 | 2.63 ( -2.19, 8.23) | 0.255 | 10 | 70 | 10,510 |
| Diuretic continuation in 180 days among prior users (%) | 83.41 | 4.88 ( -3.86, 13.82) | 0.270 | 23 | 81 | 2,913 | 78.98 | 5.39 ( -0.25, 11.03) | 0.061 | 14 | 85 | 7,830 |

**B**

| | Most deprived quintile | | | | | | Least deprived quintile | | | | | |
|---|---|---|---|---|---|---|---|---|---|---|---|---|
| | Mean below threshold | Effect of AKI alert | | Bandwidth (micromol/L) | | Effective sample size | Mean below threshold | Effect of AKI alert | | Bandwidth (micromol/L) | | Effective sample size |
| | | Estimate (95% CI) | P value | below | above | | | Estimate (95% CI) | P value | below | above | |
| ***Primary outcome*** | | | | | | | | | | | | |
| Death in 365 days (%) | 25.64 | 0.45 ( -6.00, 5.57) | 0.942 | 8 | 55 | 10,113 | 33.08 | 0.16 ( -7.75, 6.77) | 0.895 | 10 | 92 | 7,483 |
| Emergency hospitalization or emergency | 55.56 | -1.17 ( -6.44, 4.49) | 0.727 | 11 | 106 | 11,060 | 48.70 | 5.28 ( -2.23, 12.74) | 0.169 | 11 | 113 | 7,409 |
| ***Secondary outcome*** | | | | | | | | | | | | |
| *Heath outcome* | | | | | | | | | | | | |
| Death in 30 days (%) | 9.14 | 2.55 ( -1.17, 5.08) | 0.22 | 13 | 54 | 13,671 | 12.74 | -0.74 ( -5.95, 2.59) | 0.441 | 16 | 66 | 11,106 |
| Emergency hospitalization or emergency department attendance in 30 days (%) | 25.60 | 2.50 ( -1.91, 7.09) | 0.259 | 12 | 137 | 12,176 | 24.53 | -1.29 ( -7.02, 4.45) | 0.66 | 13 | 119 | 8,612 |
| *Kidney recovery outcome* | | | | | | | | | | | | |
| Peak inpatient creatinine after test (micromol/L) | 103.72 | 3.71 ( -6.96, 11.16) | 0.65 | 11 | 96 | 9,579 | 109.95 | 3.29 ( -9.55, 12.78) | 0.777 | 12 | 93 | 6,675 |
| Updated eGFR at 90 days (ml/min/1.73m$^2$) | 83.48 | -1.40 ( -4.93, 3.19) | 0.674 | 8 | 56 | 7,197 | 75.94 | -3.06 ( -7.03, 2.30) | 0.321 | 9 | 81 | 5,086 |
| *Care process outcome* | | | | | | | | | | | | |
| Length of inpatient stay >=7 days (%) | 44.06 | 2.07 ( -5.09, 7.62) | 0.697 | 9 | 62 | 9,651 | 52.30 | -4.46 (-13.48, 2.24) | 0.161 | 10 | 71 | 7,189 |
| Having an inpatient creatinine retest (%) | 68.41 | 0.60 ( -6.21, 5.54) | 0.912 | 8 | 56 | 9,458 | 72.57 | 7.38 ( -0.85, 13.67) | 0.083 | 9 | 62 | 6,604 |
| Proteinuria check in 365 days (%) | 21.89 | 3.13 ( -2.19, 7.29) | 0.292 | 12 | 84 | 10,063 | 22.05 | 3.39 ( -3.78, 9.35) | 0.405 | 13 | 99 | 5,991 |
| Creatinine test in 90 days (%) | 56.34 | -2.21 ( -9.19, 3.12) | 0.334 | 10 | 77 | 8,943 | 61.55 | 0.30 ( -8.55, 7.39) | 0.887 | 11 | 83 | 5,882 |
| Blood pressure measurement in 90 days (%) | 34.45 | 0.90 ( -4.35, 5.84) | 0.775 | 12 | 126 | 11,878 | 34.49 | 4.72 ( -2.31, 11.74) | 0.188 | 12 | 108 | 7,035 |
| AKI coding in primary care in 90 days (%) | 3.75 | 1.69 ( -0.79, 2.35) | 0.329 | 32 | 48 | 68,213 | 3.43 | 0.70 ( -2.24, 1.70) | 0.789 | 32 | 60 | 41,180 |
| AKI coding for hospitalization (%) | 15.38 | 8.59 ( 2.92, 10.78) | <0.001*** | 17 | 39 | 16,585 | 19.34 | 5.98 ( -1.60, 8.39) | 0.182 | 20 | 47 | 14,732 |
| Any PPI prescription in 365 days (%) | 49.45 | -3.64 ( -9.27, 1.35) | 0.144 | 13 | 126 | 11,245 | 39.55 | 6.58 ( -0.93, 14.08) | 0.086 | 13 | 113 | 6,046 |
| Any NSAID prescription in 365 days (%) | 13.07 | -3.38 ( -7.02, 0.73) | 0.112 | 11 | 152 | 8,919 | 8.30 | -0.69 ( -4.81, 3.67) | 0.791 | 12 | 136 | 5,671 |
| RAASI continuation in 180 days among prior users (%) | 79.96 | 5.89 ( -1.61, 14.32) | 0.118 | 15 | 103 | 4,089 | 79.42 | 8.56 ( 0.22, 19.28) | 0.045* | 16 | 72 | 2,554 |
| Diuretic continuation in 180 days among prior users (%) | 77.96 | 11.34 ( 2.89, 20.68) | 0.009** | 19 | 84 | 3,120 | 76.10 | 10.26 ( -0.67, 23.09) | 0.064 | 20 | 78 | 1,890 |

**C**

| | Rural | | | | | | Urban | | | | | |
|---|---|---|---|---|---|---|---|---|---|---|---|---|
| | Mean below threshold | Effect of AKI alert | | Bandwidth (micromol/L) | | Effective sample size | Mean below threshold | Effect of AKI alert | | Bandwidth (micromol/L) | | Effective sample size |
| | | Estimate (95% CI) | P value | below | above | | | Estimate (95% CI) | P value | below | above | |
| ***Primary outcome*** | | | | | | | | | | | | |
| Death in 365 days (%) | 31.87 | -0.34 ( -6.37, 4.28) | 0.701 | 8 | 64 | 14,387 | 28.77 | 1.39 ( -3.58, 5.41) | 0.69 | 6 | 49 | 23,459 |
| Emergency hospitalization or emergency | 51.67 | 0.79 ( -4.46, 5.88) | 0.788 | 9 | 123 | 15,056 | 53.63 | 0.04 ( -4.28, 4.57) | 0.948 | 7 | 95 | 23,907 |
| ***Secondary outcome*** | | | | | | | | | | | | |
| *Heath outcome* | | | | | | | | | | | | |
| Death in 30 days (%) | 13.56 | -2.27 ( -5.96, -0.05) | 0.046* | 13 | 59 | 21,944 | 10.69 | 2.25 ( -0.78, 4.57) | 0.164 | 7 | 48 | 26,708 |
| Emergency hospitalization or emergency department attendance in 30 days (%) | 27.27 | -2.98 ( -7.02, 1.04) | 0.145 | 12 | 115 | 17,324 | 25.36 | 1.28 ( -2.16, 4.45) | 0.496 | 9 | 122 | 28,034 |
| *Kidney recovery outcome* | | | | | | | | | | | | |
| Peak inpatient creatinine after test (micromol/L) | 105.89 | 3.69 ( -6.17, 9.78) | 0.658 | 10 | 76 | 13,312 | 107.27 | 6.63 ( -1.22, 12.39) | 0.108 | 7 | 93 | 24,092 |
| Updated eGFR at 90 days (ml/min/1.73m$^2$) | 79.58 | -2.31 ( -5.42, 1.95) | 0.357 | 8 | 51 | 9,593 | 80.27 | -0.19 ( -2.90, 3.16) | 0.934 | 6 | 48 | 16,550 |
| *Care process outcome* | | | | | | | | | | | | |
| Length of inpatient stay >=7 days (%) | 47.84 | 3.07 ( -3.32, 8.06) | 0.415 | 8 | 68 | 13,445 | 46.89 | -4.71 (-10.40, -0.36) | 0.036* | 6 | 53 | 22,254 |
| Having an inpatient creatinine retest (%) | 72.32 | 7.03 ( 1.19, 11.41) | 0.016* | 7 | 69 | 14,592 | 70.26 | -1.41 ( -6.50, 2.32) | 0.353 | 5 | 47 | 23,197 |
| Proteinuria check in 365 days (%) | 21.17 | 1.73 ( -3.20, 5.65) | 0.588 | 11 | 95 | 12,941 | 22.17 | 5.55 ( 1.65, 8.78) | 0.004** | 8 | 96 | 20,806 |
| Creatinine test in 90 days (%) | 58.08 | 4.05 ( -2.03, 9.12) | 0.213 | 9 | 127 | 12,412 | 57.94 | 0.85 ( -4.52, 5.07) | 0.912 | 7 | 74 | 20,452 |
| Blood pressure measurement in 90 days (%) | 34.99 | 3.26 ( -1.66, 7.81) | 0.203 | 11 | 177 | 14,592 | 35.58 | 3.62 ( -0.05, 7.09) | 0.054 | 9 | 174 | 27,447 |
| AKI coding in primary care in 90 days (%) | 3.07 | 1.78 ( -0.20, 2.47) | 0.097 | 27 | 67 | 60,527 | 3.42 | 1.02 ( -0.59, 1.62) | 0.358 | 21 | 47 | 70,816 |
| AKI coding for hospitalization (%) | 14.65 | 10.09 ( 5.06, 12.00) | <0.001*** | 16 | 38 | 23,717 | 16.79 | 4.63 ( 0.40, 6.52) | 0.027* | 11 | 32 | 26,761 |
| Any PPI prescription in 365 days (%) | 43.19 | -0.68 ( -6.29, 4.56) | 0.755 | 10 | 107 | 12,054 | 45.81 | 2.63 ( -1.61, 6.34) | 0.243 | 9 | 145 | 23,236 |
| Any NSAID prescription in 365 days (%) | 8.99 | 0.15 ( -2.75, 3.61) | 0.791 | 10 | 86 | 11,818 | 10.36 | 0.16 ( -2.13, 2.83) | 0.784 | 9 | 114 | 21,180 |
| RAASI continuation in 180 days among prior users (%) | 82.38 | -2.69 ( -8.87, 4.78) | 0.557 | 14 | 73 | 4,778 | 80.30 | 2.94 ( -1.94, 8.68) | 0.214 | 11 | 75 | 9,523 |
| Diuretic continuation in 180 days among prior users (%) | 80.93 | 2.09 ( -4.12, 8.68) | 0.485 | 24 | 93 | 5,969 | 79.10 | 6.43 ( 0.29, 12.71) | 0.040* | 14 | 89 | 6,335 |

**D**

| | Male | | | | | | Female | | | | | |
|---|---|---|---|---|---|---|---|---|---|---|---|---|
| | Mean below threshold | Effect of AKI alert | | Bandwidth (micromol/L) | | Effective sample size | Mean below threshold | Effect of AKI alert | | Bandwidth (micromol/L) | | Effective sample size |
| | | Estimate (95% CI) | P value | below | above | | | Estimate (95% CI) | P value | below | above | |
| ***Primary outcome*** | | | | | | | | | | | | |
| Death in 365 days (%) | 33.39 | -0.51 ( -6.17, 4.02) | 0.678 | 8 | 66 | 18,981 | 27.50 | 1.63 ( -3.68, 5.86) | 0.655 | 5 | 48 | 19,564 |
| Emergency hospitalization or emergency department attendance in 180 days (%) | 54.71 | -0.99 ( -5.92, 4.18) | 0.736 | 9 | 109 | 18,318 | 51.88 | 0.83 ( -4.00, 5.82) | 0.718 | 6 | 99 | 19,899 |
| ***Secondary outcome*** | | | | | | | | | | | | |
| *Heath outcome* | | | | | | | | | | | | |
| Death in 30 days (%) | 13.31 | -0.26 ( -3.86, 2.35) | 0.634 | 11 | 56 | 21,962 | 10.59 | 1.46 ( -1.84, 4.09) | 0.457 | 6 | 53 | 21,382 |
| Emergency hospitalization or emergency department attendance in 30 days (%) | 28.43 | -2.21 ( -5.97, 1.66) | 0.268 | 12 | 112 | 21,948 | 24.46 | 0.89 ( -2.83, 4.42) | 0.667 | 8 | 112 | 23,381 |
| *Kidney recovery outcome* | | | | | | | | | | | | |
| Peak inpatient creatinine after test (micromol/L) | 120.76 | 9.59 ( 0.10, 16.19) | 0.047* | 9 | 87 | 18,235 | 97.05 | 0.34 ( -7.67, 6.20) | 0.836 | 7 | 87 | 18,339 |
| Updated eGFR at 90 days (ml/min/1.73m$^2$) | 81.84 | -3.30 ( -6.14, 0.40) | 0.085 | 8 | 47 | 12,502 | 78.85 | 1.02 ( -1.97, 4.78) | 0.415 | 5 | 47 | 13,568 |
| *Care process outcome* | | | | | | | | | | | | |
| Length of inpatient stay >=7 days (%) | 46.58 | 0.13 ( -5.98, 5.07) | 0.871 | 8 | 62 | 17,388 | 47.61 | -3.34 ( -9.61, 1.26) | 0.132 | 5 | 52 | 18,434 |
| Having an inpatient creatinine retest (%) | 74.17 | 4.91 ( -0.47, 9.19) | 0.077 | 7 | 73 | 18,436 | 68.87 | -1.07 ( -6.82, 2.79) | 0.412 | 5 | 42 | 18,981 |
| Proteinuria check in 365 days (%) | 22.30 | 2.76 ( -1.85, 6.37) | 0.281 | 12 | 99 | 16,053 | 21.57 | 5.46 ( 1.07, 9.29) | 0.014* | 7 | 81 | 16,208 |
| Creatinine test in 90 days (%) | 62.21 | -1.14 ( -7.08, 3.57) | 0.518 | 8 | 89 | 15,632 | 55.35 | 4.51 ( -1.58, 9.64) | 0.159 | 5 | 90 | 16,318 |
| Blood pressure measurement in 90 days (%) | 36.16 | 2.06 ( -2.39, 6.15) | 0.389 | 11 | 185 | 20,133 | 34.90 | 4.64 ( 0.34, 8.82) | 0.034* | 8 | 113 | 19,336 |
| AKI coding in primary care in 90 days (%) | 4.24 | 1.46 ( -0.61, 1.94) | 0.309 | 32 | 57 | 87,617 | 2.72 | 1.17 ( -0.40, 1.97) | 0.195 | 17 | 49 | 40,783 |
| AKI coding for hospitalization (%) | 19.42 | 9.80 ( 4.85, 12.11) | <0.001*** | 14 | 41 | 21,896 | 13.97 | 3.79 ( -0.58, 5.70) | 0.11 | 10 | 30 | 23,552 |
| Any PPI prescription in 365 days (%) | 45.29 | 2.06 ( -2.83, 6.80) | 0.419 | 11 | 128 | 16,478 | 44.75 | 0.10 ( -4.62, 4.13) | 0.913 | 9 | 91 | 17,835 |
| Any NSAID prescription in 365 days (%) | 7.83 | 0.54 ( -1.90, 3.13) | 0.634 | 12 | 124 | 16,418 | 11.15 | 0.40 ( -2.59, 3.67) | 0.735 | 7 | 86 | 15,045 |
| RAASI continuation in 180 days among prior users (%) | 81.56 | 2.56 ( -2.83, 9.08) | 0.303 | 13 | 73 | 7,122 | 80.47 | 0.60 ( -5.22, 7.14) | 0.76 | 10 | 75 | 6,858 |
| Diuretic continuation in 180 days among prior users (%) | 80.91 | 3.21 ( -3.05, 9.57) | 0.312 | 21 | 84 | 5,488 | 79.04 | 5.45 ( -0.99, 11.98) | 0.097 | 14 | 92 | 5,732 |

**E**

| | With major surgery during admission | | | | | | Without major surgery during admission | | | | | |
|---|---|---|---|---|---|---|---|---|---|---|---|---|
| | Mean below threshold | Effect of AKI alert | | Bandwidth (micromol/L) | | Effective sample size | Mean below threshold | Effect of AKI alert | | Bandwidth (micromol/L) | | Effective sample size |
| | | Estimate (95% CI) | P value | below | above | | | Estimate (95% CI) | P value | below | above | |
| ***Primary outcome*** | | | | | | | | | | | | |
| Death in 365 days (%) | 26.74 | 10.14 (-10.73, 27.76) | 0.386 | 23 | 107 | 1,223 | 29.84 | 1.28 ( -3.12, 4.73) | 0.688 | 5 | 46 | 34,332 |
| Emergency hospitalization or emergency department attendance in 180 days (%) | 41.56 | 3.85 (-21.37, 28.03) | 0.792 | 24 | 68 | 1,160 | 53.08 | 0.01 ( -3.94, 4.10) | 0.968 | 6 | 98 | 33,534 |
| ***Secondary outcome*** | | | | | | | | | | | | |
| *Heath outcome* | | | | | | | | | | | | |
| Death in 30 days (%) | 9.30 | 6.23 ( -7.58, 19.44) | 0.39 | 27 | 77 | 1,624 | 11.68 | 0.32 ( -2.45, 2.42) | 0.991 | 7 | 45 | 36,744 |
| Emergency hospitalization or emergency department attendance in 30 days (%) | 18.18 | -0.90 (-21.88, 19.29) | 0.902 | 26 | 65 | 1,269 | 26.07 | -0.42 ( -3.39, 2.48) | 0.761 | 8 | 119 | 38,951 |
| *Kidney recovery outcome* | | | | | | | | | | | | |
| Peak inpatient creatinine after test (micromol/L) | 132.36 | 16.90 (-22.49, 34.64) | 0.677 | 37 | 87 | 2,938 | 106.48 | 6.85 ( -0.02, 11.59) | 0.051 | 7 | 84 | 33,564 |
| Updated eGFR at 90 days (ml/min/1.73m$^2$) | 82.30 | 12.30 ( 1.23, 30.54) | 0.034* | 21 | 56 | 799 | 80.02 | -0.54 ( -2.90, 2.41) | 0.857 | 5 | 41 | 23,394 |
| *Care process outcome* | | | | | | | | | | | | |
| Length of inpatient stay >=7 days (%) | 96.34 | 2.84 ( -6.11, 10.95) | 0.578 | 20 | 75 | 991 | 46.75 | -3.02 ( -8.05, 0.86) | 0.114 | 5 | 49 | 32,448 |
| Having an inpatient creatinine retest (%) | 96.51 | 2.10 ( -6.39, 10.27) | 0.648 | 23 | 76 | 1,283 | 70.72 | 0.69 ( -3.94, 4.13) | 0.963 | 5 | 44 | 31,616 |
| Proteinuria check in 365 days (%) | 27.87 | -6.18 (-28.84, 11.21) | 0.388 | 36 | 69 | 2,126 | 21.78 | 4.65 ( 1.07, 7.62) | 0.009** | 7 | 85 | 28,214 |
| Creatinine test in 90 days (%) | 71.43 | -27.00 (-55.49, -8.76) | 0.007** | 29 | 65 | 1,585 | 57.86 | 1.92 ( -2.90, 5.78) | 0.516 | 6 | 76 | 28,412 |
| Blood pressure measurement in 90 days (%) | 34.29 | -4.22 (-24.46, 14.24) | 0.605 | 33 | 97 | 1,985 | 35.39 | 3.78 ( 0.57, 6.79) | 0.020* | 9 | 153 | 37,622 |
| AKI coding in primary care in 90 days (%) | 4.23 | -2.36 ( -6.18, 0.89) | 0.142 | 39 | 40 | 2,760 | 3.29 | 0.77 ( -0.63, 1.35) | 0.473 | 17 | 44 | 71,463 |
| AKI coding for hospitalization (%) | 14.29 | 14.97 ( -3.14, 32.17) | 0.107 | 34 | 65 | 2,220 | 16.09 | 5.79 ( 2.11, 7.48) | <0.001*** | 10 | 29 | 35,830 |
| Any PPI prescription in 365 days (%) | 52.46 | -2.93 (-31.15, 22.80) | 0.761 | 25 | 82 | 1,064 | 44.88 | 2.12 ( -1.62, 5.45) | 0.288 | 8 | 129 | 31,689 |
| Any NSAID prescription in 365 days (%) | 9.84 | -10.92 (-25.27, 2.18) | 0.099 | 27 | 75 | 1,232 | 9.91 | 0.33 ( -1.63, 2.63) | 0.646 | 8 | 98 | 28,722 |
| RAASI continuation in 180 days among prior users (%) | 76.67 | 9.09 (-13.69, 39.54) | 0.341 | 35 | 107 | 636 | 80.99 | 2.29 ( -2.12, 7.42) | 0.277 | 10 | 68 | 11,804 |
| Diuretic continuation in 180 days among prior users (%) | 75.00 | -16.43 (-65.22, 19.64) | 0.292 | 50 | 63 | 485 | 79.76 | 4.93 ( -0.10, 10.01) | 0.055 | 14 | 81 | 9,211 |

**F**

| | With history of vascular disease | | | | | | Without history of vascular disease | | | | | |
|---|---|---|---|---|---|---|---|---|---|---|---|---|
| | Mean below threshold | Effect of AKI alert | | Bandwidth (micromol/L) | | Effective sample size | Mean below threshold | Effect of AKI alert | | Bandwidth (micromol/L) | | Effective sample size |
| | | Estimate (95% CI) | P value | below | above | | | Estimate (95% CI) | P value | below | above | |
| ***Primary outcome*** | | | | | | | | | | | | |
| Death in 365 days (%) | 39.53 | 4.22 ( -1.90, 9.13) | 0.199 | 8 | 69 | 16,796 | 24.85 | -1.90 ( -6.70, 1.91) | 0.276 | 6 | 49 | 22,229 |
| Emergency hospitalization or emergency department attendance in 180 days (%) | 57.83 | 0.82 ( -4.13, 5.85) | 0.735 | 10 | 135 | 16,419 | 50.72 | -0.11 ( -4.28, 4.28) | 1 | 7 | 104 | 25,400 |
| ***Secondary outcome*** | | | | | | | | | | | | |
| *Heath outcome* | | | | | | | | | | | | |
| Death in 30 days (%) | 15.92 | 1.23 ( -2.95, 4.20) | 0.733 | 11 | 61 | 19,798 | 9.48 | 0.13 ( -2.66, 2.20) | 0.855 | 8 | 48 | 25,620 |
| Emergency hospitalization or emergency department attendance in 30 days (%) | 28.67 | -1.64 ( -6.40, 2.37) | 0.367 | 12 | 104 | 17,042 | 24.75 | -0.08 ( -3.19, 3.14) | 0.989 | 9 | 120 | 27,644 |
| *Kidney recovery outcome* | | | | | | | | | | | | |
| Peak inpatient creatinine after test (micromol/L) | 122.21 | 7.50 ( -2.34, 14.43) | 0.158 | 10 | 89 | 15,709 | 97.46 | 3.88 ( -3.65, 8.81) | 0.417 | 8 | 81 | 21,569 |
| Updated eGFR at 90 days (ml/min/1.73m$^2$) | 69.40 | -2.53 ( -5.58, 1.15) | 0.197 | 8 | 48 | 9,921 | 85.45 | -0.02 ( -2.60, 3.35) | 0.805 | 6 | 47 | 15,841 |
| *Care process outcome* | | | | | | | | | | | | |
| Length of inpatient stay >=7 days (%) | 57.20 | 4.03 ( -2.28, 8.96) | 0.244 | 8 | 79 | 15,583 | 42.29 | -5.38 (-11.00, -1.00) | 0.019* | 6 | 50 | 21,161 |
| Having an inpatient creatinine retest (%) | 79.19 | 4.40 ( -0.74, 7.96) | 0.103 | 8 | 99 | 17,003 | 66.75 | 0.12 ( -5.37, 4.28) | 0.824 | 5 | 43 | 21,483 |
| Proteinuria check in 365 days (%) | 26.21 | 3.74 ( -1.30, 7.98) | 0.159 | 13 | 127 | 12,898 | 20.06 | 3.56 ( -0.36, 6.86) | 0.078 | 8 | 89 | 19,730 |
| Creatinine test in 90 days (%) | 67.18 | -0.60 ( -6.48, 3.87) | 0.621 | 11 | 106 | 13,840 | 53.86 | 1.12 ( -4.13, 5.38) | 0.796 | 6 | 76 | 20,355 |
| Blood pressure measurement in 90 days (%) | 43.92 | 3.27 ( -1.22, 8.15) | 0.147 | 14 | 88 | 17,790 | 31.56 | 2.84 ( -0.78, 6.11) | 0.13 | 9 | 134 | 27,036 |
| AKI coding in primary care in 90 days (%) | 5.03 | 2.20 ( -0.38, 3.18) | 0.123 | 26 | 60 | 41,333 | 2.52 | 0.71 ( -0.68, 1.24) | 0.568 | 21 | 48 | 68,995 |
| AKI coding for hospitalization (%) | 24.87 | 8.03 ( 1.88, 9.95) | 0.004** | 17 | 42 | 23,597 | 11.98 | 6.50 ( 2.82, 8.18) | <0.001*** | 11 | 30 | 26,722 |
| Any PPI prescription in 365 days (%) | 55.43 | 0.96 ( -3.84, 5.94) | 0.673 | 15 | 121 | 14,757 | 40.67 | 0.73 ( -3.09, 4.19) | 0.767 | 10 | 106 | 25,334 |
| Any NSAID prescription in 365 days (%) | 4.32 | 0.74 ( -1.31, 2.82) | 0.473 | 13 | 130 | 12,911 | 12.19 | 0.11 ( -2.34, 3.02) | 0.805 | 9 | 83 | 21,119 |
| RAASI continuation in 180 days among prior users (%) | 81.03 | 2.66 ( -2.99, 9.37) | 0.312 | 13 | 65 | 7,030 | 80.84 | -0.46 ( -5.71, 5.73) | 0.997 | 11 | 82 | 7,080 |
| Diuretic continuation in 180 days among prior users (%) | 83.17 | 5.74 ( 0.77, 11.24) | 0.025* | 17 | 75 | 7,312 | 73.32 | 3.83 ( -4.07, 11.51) | 0.349 | 18 | 119 | 4,692 |

**G**

| | With history of diabetes | | | | | | Without history of diabetes | | | | | |
|---|---|---|---|---|---|---|---|---|---|---|---|---|
| | Mean below threshold | Effect of AKI alert | | Bandwidth (micromol/L) | | Effective sample size | Mean below threshold | Effect of AKI alert | | Bandwidth (micromol/L) | | Effective sample size |
| | | Estimate (95% CI) | P value | below | above | | | Estimate (95% CI) | P value | below | above | |
| ***Primary outcome*** | | | | | | | | | | | | |
| Death in 365 days (%) | 30.81 | -0.48 ( -6.38, 4.04) | 0.659 | 10 | 73 | 15,833 | 29.43 | 0.24 ( -4.67, 4.09) | 0.896 | 6 | 45 | 23,950 |
| Emergency hospitalization or emergency department attendance in 180 days (%) | 55.32 | 0.84 ( -4.46, 6.21) | 0.748 | 11 | 90 | 14,853 | 52.08 | 0.62 ( -3.50, 4.82) | 0.756 | 7 | 116 | 25,270 |
| ***Secondary outcome*** | | | | | | | | | | | | |
| *Heath outcome* | | | | | | | | | | | | |
| Death in 30 days (%) | 11.03 | 0.85 ( -2.77, 3.16) | 0.897 | 15 | 60 | 21,150 | 11.90 | 0.60 ( -2.57, 3.02) | 0.876 | 7 | 47 | 26,018 |
| Emergency hospitalization or emergency department attendance in 30 days (%) | 25.69 | 1.77 ( -2.69, 6.37) | 0.426 | 12 | 105 | 16,056 | 26.11 | -1.56 ( -4.77, 1.45) | 0.294 | 9 | 127 | 29,306 |
| *Kidney recovery outcome* | | | | | | | | | | | | |
| Peak inpatient creatinine after test (micromol/L) | 118.93 | 0.26 (-11.21, 8.33) | 0.773 | 11 | 94 | 14,318 | 101.70 | 9.31 ( 2.53, 13.96) | 0.005** | 7 | 80 | 24,017 |
| Updated eGFR at 90 days (ml/min/1.73m$^2$) | 74.21 | -2.01 ( -5.35, 2.18) | 0.41 | 9 | 65 | 10,259 | 82.48 | -0.02 ( -2.54, 3.28) | 0.804 | 6 | 43 | 16,235 |
| *Care process outcome* | | | | | | | | | | | | |
| Length of inpatient stay >=7 days (%) | 49.31 | 0.88 ( -5.31, 5.52) | 0.97 | 11 | 96 | 15,418 | 46.40 | -2.17 ( -7.69, 2.13) | 0.267 | 5 | 45 | 22,091 |
| Having an inpatient creatinine retest (%) | 76.05 | 4.74 ( -1.10, 8.51) | 0.13 | 9 | 98 | 14,864 | 69.01 | 0.32 ( -4.75, 4.18) | 0.901 | 5 | 43 | 23,644 |
| Proteinuria check in 365 days (%) | 43.18 | 3.25 ( -2.15, 8.01) | 0.258 | 15 | 134 | 14,175 | 13.85 | 2.65 ( -0.29, 5.19) | 0.079 | 9 | 115 | 22,054 |
| Creatinine test in 90 days (%) | 65.05 | 1.61 ( -4.25, 6.24) | 0.71 | 12 | 114 | 13,573 | 55.26 | 1.90 ( -3.21, 6.02) | 0.552 | 6 | 73 | 20,954 |
| Blood pressure measurement in 90 days (%) | 41.96 | 2.33 ( -1.83, 6.36) | 0.278 | 19 | 132 | 21,819 | 32.85 | 3.70 ( 0.21, 7.15) | 0.037* | 9 | 136 | 26,073 |
| AKI coding in primary care in 90 days (%) | 5.08 | 2.83 ( -0.06, 3.20) | 0.059 | 36 | 65 | 73,407 | 2.62 | 0.76 ( -0.59, 1.42) | 0.422 | 18 | 48 | 53,579 |
| AKI coding for hospitalization (%) | 23.96 | 8.40 ( 1.85, 9.69) | 0.004** | 21 | 43 | 25,755 | 13.07 | 6.72 ( 2.97, 8.53) | <0.001*** | 10 | 30 | 28,496 |
| Any PPI prescription in 365 days (%) | 50.14 | 1.02 ( -3.99, 5.85) | 0.711 | 15 | 180 | 15,415 | 43.01 | 0.59 ( -3.47, 4.18) | 0.855 | 9 | 119 | 23,881 |
| Any NSAID prescription in 365 days (%) | 7.97 | 0.87 ( -1.98, 4.08) | 0.497 | 12 | 115 | 11,544 | 10.64 | -0.20 ( -2.48, 2.47) | 0.997 | 8 | 101 | 21,770 |
| RAASI continuation in 180 days among prior users (%) | 82.22 | 1.33 ( -4.24, 7.71) | 0.569 | 14 | 75 | 6,763 | 79.98 | 1.62 ( -3.68, 7.96) | 0.471 | 11 | 80 | 7,276 |
| Diuretic continuation in 180 days among prior users (%) | 81.03 | 5.00 ( -1.82, 12.30) | 0.146 | 18 | 104 | 5,014 | 78.85 | 4.51 ( -1.30, 10.43) | 0.127 | 15 | 87 | 6,479 |

**Table S5: Effects of AKI alerts in emergency hospitalization setting by (A) alert approach, (B) resident neighbourhood measures of deprivation (WIMD), (C) rurality, (D) sex, (E) presence of major surgery during the admission, (F) history of vascular disease and (G) diabetes.**

**A**

| | Most deprived quintile | | | | | | Least deprived quintile | | | | | |
|---|---|---|---|---|---|---|---|---|---|---|---|---|
| | Mean below threshold | Effect of AKI alert | | Bandwidth (micromol/L) | | Effective sample size | Mean below threshold | Effect of AKI alert | | Bandwidth (micromol/L) | | Effective sample size |
| | | Estimate (95% CI) | P value | below | above | | | Estimate (95% CI) | P value | below | above | |
| ***Primary outcome*** | | | | | | | | | | | | |
| Death in 365 days (%) | 26.13 | -3.43 (-12.59, 3.56) | 0.273 | 11 | 67 | 3,558 | 26.10 | 6.54 ( -2.26, 14.26) | 0.155 | 15 | 86 | 3,758 |
| Emergency hospitalization or emergency | 64.41 | 3.94 ( -4.17, 10.46) | 0.399 | 13 | 118 | 4,511 | 59.43 | -0.90 (-11.12, 6.95) | 0.652 | 15 | 87 | 3,333 |
| ***Secondary outcome*** | | | | | | | | | | | | |
| *Heath outcome* | | | | | | | | | | | | |
| Death in 30 days (%) | 6.91 | -0.04 ( -4.87, 3.44) | 0.736 | 18 | 59 | 6,892 | 6.80 | 4.17 ( -1.53, 7.91) | 0.185 | 22 | 68 | 8,332 |
| Emergency hospitalization or emergency department attendance in 30 days (%) | 45.05 | 0.47 ( -9.54, 6.97) | 0.761 | 13 | 61 | 4,405 | 40.13 | 1.10 ( -8.95, 8.24) | 0.935 | 17 | 86 | 4,711 |
| *Kidney recovery outcome* | | | | | | | | | | | | |
| Updated eGFR at 90 days (ml/min/1.73m2) | 84.76 | 1.96 ( -3.18, 9.20) | 0.34 | 11 | 54 | 2,121 | 79.06 | -2.00 ( -7.40, 6.34) | 0.88 | 11 | 41 | 1,599 |
| *Care process outcome* | | | | | | | | | | | | |
| Proteinuria check in 365 days (%) | 22.76 | 1.72 ( -6.36, 8.24) | 0.801 | 16 | 76 | 4,787 | 21.66 | -0.78 (-12.06, 7.22) | 0.623 | 15 | 49 | 2,813 |
| Creatinine test in 90 days (%) | 66.32 | -1.11 (-10.44, 6.12) | 0.609 | 13 | 86 | 3,866 | 70.10 | 1.10 (-10.35, 9.06) | 0.897 | 15 | 67 | 2,884 |
| Blood pressure measurement in 90 days (%) | 36.63 | -1.29 ( -9.05, 5.31) | 0.609 | 17 | 88 | 6,201 | 37.94 | -0.24 (-10.31, 9.89) | 0.968 | 15 | 75 | 2,890 |
| AKI coding in primary care in 90 days (%) | 2.26 | 2.71 ( -0.27, 5.07) | 0.078 | 26 | 81 | 15,626 | 2.01 | 6.59 ( 2.43, 9.72) | 0.001** | 31 | 71 | 19,103 |
| Any PPI prescription in 365 days (%) | 53.86 | 0.61 ( -7.01, 8.75) | 0.829 | 19 | 70 | 6,906 | 41.84 | 1.40 ( -8.80, 12.11) | 0.756 | 17 | 79 | 3,206 |
| Any NSAID prescription in 365 days (%) | 11.18 | 0.38 ( -5.41, 6.67) | 0.838 | 13 | 80 | 2,965 | 8.01 | 2.95 ( -2.42, 10.34) | 0.224 | 16 | 58 | 2,827 |
| RAASI continuation in 180 days among prior users (%) | 86.31 | -4.91 (-14.29, 6.46) | 0.459 | 18 | 71 | 1,541 | 87.63 | 3.99 ( -5.47, 16.10) | 0.334 | 20 | 93 | 1,540 |
| Diuretic continuation in 180 days among prior users (%) | 83.76 | 3.91 ( -8.46, 15.53) | 0.564 | 23 | 72 | 1,940 | 85.07 | 0.59 (-12.36, 12.34) | 0.999 | 31 | 108 | 2,309 |

**B**

| | Rural | | | | | | Urban | | | | | |
|---|---|---|---|---|---|---|---|---|---|---|---|---|
| | Mean below threshold | Effect of AKI alert | | Bandwidth (micromol/L) | | Effective sample size | Mean below threshold | Effect of AKI alert | | Bandwidth (micromol/L) | | Effective sample size |
| | | Estimate (95% CI) | P value | below | above | | | Estimate (95% CI) | P value | below | above | |
| ***Primary outcome*** | | | | | | | | | | | | |
| Death in 365 days (%) | 26.17 | 0.38 ( -6.60, 5.75) | 0.893 | 12 | 76 | 6,119 | 24.86 | 2.42 ( -3.96, 7.75) | 0.525 | 7 | 70 | 7,002 |
| Emergency hospitalization or emergency department attendance in 180 days (%) | 58.49 | 4.79 ( -2.82, 10.02) | 0.271 | 13 | 85 | 6,150 | 62.61 | 3.66 ( -3.04, 8.50) | 0.353 | 9 | 68 | 7,693 |
| ***Secondary outcome*** | | | | | | | | | | | | |
| *Heath outcome* | | | | | | | | | | | | |
| Death in 30 days (%) | 6.52 | 0.21 ( -3.32, 2.61) | 0.814 | 20 | 65 | 13,384 | 6.70 | 0.86 ( -2.26, 2.88) | 0.815 | 14 | 56 | 14,391 |
| Emergency hospitalization or emergency department attendance in 30 days (%) | 37.20 | 5.46 ( -2.25, 10.42) | 0.206 | 14 | 71 | 6,814 | 41.70 | 2.07 ( -4.97, 6.80) | 0.761 | 9 | 50 | 8,276 |
| *Kidney recovery outcome* | | | | | | | | | | | | |
| Updated eGFR at 90 days (ml/min/1.73m2) | 82.31 | -1.99 ( -5.91, 4.23) | 0.746 | 11 | 44 | 3,507 | 81.11 | 1.14 ( -2.82, 6.45) | 0.442 | 7 | 45 | 4,138 |
| *Care process outcome* | | | | | | | | | | | | |
| Proteinuria check in 365 days (%) | 21.17 | 3.70 ( -2.70, 9.12) | 0.287 | 16 | 87 | 7,299 | 21.67 | 0.39 ( -5.44, 5.02) | 0.937 | 11 | 70 | 6,889 |
| Creatinine test in 90 days (%) | 63.36 | 9.56 ( 2.29, 15.71) | 0.009** | 12 | 102 | 5,292 | 67.85 | -2.27 ( -9.60, 2.70) | 0.272 | 9 | 58 | 6,360 |
| Blood pressure measurement in 90 days (%) | 35.99 | 4.45 ( -2.59, 10.32) | 0.241 | 15 | 79 | 6,647 | 36.87 | -2.94 ( -8.88, 1.76) | 0.19 | 12 | 75 | 9,019 |
| AKI coding in primary care in 90 days (%) | 1.20 | 3.70 ( 1.21, 5.68) | 0.002** | 26 | 70 | 23,252 | 2.39 | 3.20 ( 1.22, 4.32) | <0.001*** | 29 | 78 | 62,094 |
| Any PPI prescription in 365 days (%) | 45.19 | -1.92 ( -9.08, 4.63) | 0.525 | 16 | 86 | 7,298 | 49.16 | -0.99 ( -6.62, 4.29) | 0.676 | 13 | 110 | 8,776 |
| Any NSAID prescription in 365 days (%) | 8.70 | 2.37 ( -1.11, 6.60) | 0.163 | 14 | 120 | 5,730 | 9.12 | 2.80 ( -0.54, 6.88) | 0.094 | 10 | 69 | 6,135 |
| RAASI continuation in 180 days among prior users (%) | 87.39 | -3.95 (-11.66, 5.22) | 0.454 | 19 | 75 | 2,616 | 85.21 | -0.64 ( -7.53, 6.41) | 0.875 | 12 | 95 | 2,863 |
| Diuretic continuation in 180 days among prior users (%) | 81.66 | 0.40 ( -9.94, 9.77) | 0.987 | 25 | 61 | 3,160 | 83.61 | 2.67 ( -4.13, 9.12) | 0.461 | 19 | 107 | 3,989 |

**C**

| | Male | | | | | | Female | | | | | |
|---|---|---|---|---|---|---|---|---|---|---|---|---|
| | Mean below threshold | Effect of AKI alert | | Bandwidth (micromol/L) | | Effective sample size | Mean below threshold | Effect of AKI alert | | Bandwidth (micromol/L) | | Effective sample size |
| | | Estimate (95% CI) | P value | below | above | | | Estimate (95% CI) | P value | below | above | |
| ***Primary outcome*** | | | | | | | | | | | | |
| Death in 365 days (%) | 30.08 | 1.54 ( -5.51, 8.15) | 0.704 | 10 | 85 | 5,524 | 22.59 | 0.13 ( -6.61, 5.79) | 0.897 | 7 | 61 | 5,662 |
| Emergency hospitalization or emergency department attendance in 180 days (%) | 64.08 | 3.50 ( -4.50, 9.14) | 0.505 | 11 | 67 | 5,427 | 59.52 | 3.32 ( -3.60, 8.95) | 0.404 | 8 | 88 | 6,471 |
| ***Secondary outcome*** | | | | | | | | | | | | |
| *Heath outcome* | | | | | | | | | | | | |
| Death in 30 days (%) | 7.57 | 1.92 ( -1.78, 4.64) | 0.383 | 17 | 77 | 10,906 | 6.10 | -0.33 ( -3.55, 1.88) | 0.547 | 12 | 52 | 9,820 |
| Emergency hospitalization or emergency department attendance in 30 days (%) | 45.03 | 1.59 ( -6.61, 6.88) | 0.969 | 12 | 57 | 6,460 | 37.35 | 3.32 ( -3.95, 8.70) | 0.462 | 8 | 56 | 7,002 |
| *Kidney recovery outcome* | | | | | | | | | | | | |
| Updated eGFR at 90 days (ml/min/1.73m2) | 82.40 | 1.38 ( -3.63, 7.69) | 0.481 | 9 | 54 | 3,041 | 80.96 | 0.09 ( -3.48, 5.45) | 0.666 | 8 | 35 | 4,080 |
| *Care process outcome* | | | | | | | | | | | | |
| Proteinuria check in 365 days (%) | 21.12 | 2.59 ( -3.94, 8.13) | 0.496 | 14 | 108 | 5,144 | 21.70 | 0.76 ( -5.20, 5.73) | 0.924 | 10 | 73 | 6,691 |
| Creatinine test in 90 days (%) | 72.09 | -2.37 (-10.96, 4.05) | 0.367 | 10 | 77 | 4,496 | 63.10 | 4.83 ( -2.64, 10.44) | 0.243 | 8 | 64 | 5,404 |
| Blood pressure measurement in 90 days (%) | 40.64 | -3.27 ( -9.50, 2.38) | 0.24 | 16 | 101 | 8,458 | 34.31 | 1.75 ( -4.44, 6.35) | 0.729 | 11 | 58 | 8,659 |
| AKI coding in primary care in 90 days (%) | 2.67 | 4.24 ( 1.77, 6.16) | <0.001*** | 30 | 76 | 35,855 | 1.59 | 2.79 ( 0.72, 4.00) | 0.005** | 25 | 61 | 41,765 |
| Any PPI prescription in 365 days (%) | 48.63 | -2.17 ( -9.35, 5.44) | 0.605 | 14 | 76 | 5,657 | 47.37 | 0.04 ( -5.97, 5.27) | 0.903 | 12 | 109 | 8,661 |
| Any NSAID prescription in 365 days (%) | 6.65 | 4.59 ( 0.92, 9.07) | 0.016* | 13 | 69 | 5,076 | 10.18 | 3.65 ( -0.02, 7.99) | 0.051 | 8 | 74 | 5,266 |
| RAASI continuation in 180 days among prior users (%) | 85.67 | -0.69 ( -7.92, 7.88) | 0.996 | 15 | 70 | 2,309 | 86.06 | -3.39 (-10.97, 3.92) | 0.354 | 13 | 98 | 2,534 |
| Diuretic continuation in 180 days among prior users (%) | 83.07 | 5.69 ( -2.70, 14.05) | 0.184 | 21 | 58 | 2,434 | 82.94 | -0.70 ( -8.64, 5.86) | 0.707 | 19 | 129 | 3,604 |

**D**

| | With history of vascular disease: Mean below threshold | With history of vascular disease: Effect of AKI alert Estimate (95% CI) | With history of vascular disease: Effect of AKI alert P value | With history of vascular disease: Bandwidth (micromol/L) below | With history of vascular disease: Bandwidth (micromol/L) above | With history of vascular disease: Effective sample size | Without history of vascular disease: Mean below threshold | Without history of vascular disease: Effect of AKI alert Estimate (95% CI) | Without history of vascular disease: Effect of AKI alert P value | Without history of vascular disease: Bandwidth (micromol/L) below | Without history of vascular disease: Bandwidth (micromol/L) above | Without history of vascular disease: Effective sample size |
|---|---|---|---|---|---|---|---|---|---|---|---|---|
| ***Primary outcome*** | | | | | | | | | | | | |
| Death in 365 days (%) | 31.00 | 3.57 ( -3.29, 10.76) | 0.297 | 11 | 77 | 4,850 | 22.73 | -0.79 ( -7.08, 4.12) | 0.605 | 8 | 59 | 6,790 |
| Emergency hospitalization or emergency department attendance in 180 days (%) | 65.71 | 1.97 ( -5.52, 8.12) | 0.709 | 11 | 89 | 5,356 | 59.12 | 4.73 ( -1.75, 9.08) | 0.185 | 10 | 63 | 8,454 |
| ***Secondary outcome*** | | | | | | | | | | | | |
| *Heath outcome* | | | | | | | | | | | | |
| Death in 30 days (%) | 7.96 | 2.51 ( -1.36, 5.74) | 0.227 | 18 | 77 | 9,321 | 6.03 | -0.90 ( -3.76, 0.88) | 0.223 | 15 | 51 | 14,957 |
| Emergency hospitalization or emergency department attendance in 30 days (%) | 43.95 | 1.62 ( -6.57, 7.83) | 0.865 | 12 | 76 | 5,302 | 38.42 | 3.35 ( -3.37, 7.55) | 0.454 | 10 | 46 | 9,225 |
| *Kidney recovery outcome* | | | | | | | | | | | | |
| Updated eGFR at 90 days (ml/min/1.73m2) | 66.89 | 1.71 ( -2.28, 7.09) | 0.314 | 10 | 50 | 3,004 | 88.91 | -1.59 ( -5.43, 3.43) | 0.659 | 7 | 46 | 4,426 |
| *Care process outcome* | | | | | | | | | | | | |
| Proteinuria check in 365 days (%) | 27.54 | -0.92 ( -8.50, 5.84) | 0.717 | 14 | 83 | 4,373 | 19.05 | 3.00 ( -2.20, 7.13) | 0.301 | 12 | 77 | 9,252 |
| Creatinine test in 90 days (%) | 74.72 | -1.64 ( -9.73, 4.06) | 0.42 | 12 | 77 | 4,364 | 62.62 | 4.25 ( -2.62, 9.34) | 0.271 | 9 | 76 | 7,375 |
| Blood pressure measurement in 90 days (%) | 47.56 | 0.20 ( -6.40, 5.88) | 0.934 | 20 | 79 | 9,769 | 31.77 | -0.20 ( -5.17, 4.18) | 0.836 | 13 | 86 | 10,630 |
| AKI coding in primary care in 90 days (%) | 2.88 | 4.56 ( 1.50, 6.83) | 0.002** | 25 | 78 | 16,468 | 1.58 | 2.64 ( 0.89, 3.81) | 0.002** | 27 | 68 | 62,326 |
| Any PPI prescription in 365 days (%) | 55.38 | -2.47 (-10.20, 5.24) | 0.529 | 15 | 77 | 5,377 | 44.72 | 0.11 ( -4.22, 5.04) | 0.863 | 17 | 74 | 17,370 |
| Any NSAID prescription in 365 days (%) | 3.69 | 1.17 ( -1.60, 4.40) | 0.361 | 15 | 86 | 4,867 | 11.12 | 3.85 ( 0.43, 8.04) | 0.029* | 10 | 70 | 7,198 |
| RAASI continuation in 180 days among prior users (%) | 86.47 | -2.95 ( -9.98, 3.70) | 0.368 | 14 | 107 | 2,652 | 85.15 | -2.00 ( -9.79, 7.32) | 0.777 | 16 | 58 | 2,575 |
| Diuretic continuation in 180 days among prior users (%) | 88.34 | 1.61 ( -4.83, 7.65) | 0.658 | 20 | 72 | 4,132 | 73.12 | 0.96 ( -8.87, 9.89) | 0.915 | 27 | 109 | 4,776 |

**E**

| | With history of diabetes: Mean below threshold | With history of diabetes: Effect of AKI alert Estimate (95% CI) | With history of diabetes: Effect of AKI alert P value | With history of diabetes: Bandwidth (micromol/L) below | With history of diabetes: Bandwidth (micromol/L) above | With history of diabetes: Effective sample size | Without history of diabetes: Mean below threshold | Without history of diabetes: Effect of AKI alert Estimate (95% CI) | Without history of diabetes: Effect of AKI alert P value | Without history of diabetes: Bandwidth (micromol/L) below | Without history of diabetes: Bandwidth (micromol/L) above | Without history of diabetes: Effective sample size |
|---|---|---|---|---|---|---|---|---|---|---|---|---|
| ***Primary outcome*** | | | | | | | | | | | | |
| Death in 365 days (%) | 28.11 | 2.55 ( -4.96, 10.35) | 0.49 | 12 | 71 | 4,144 | 24.39 | 0.80 ( -5.20, 5.55) | 0.949 | 7 | 59 | 7,556 |
| Emergency hospitalization or emergency department attendance in 180 days (%) | 67.87 | 2.89 ( -6.21, 9.78) | 0.662 | 11 | 73 | 4,153 | 58.98 | 4.24 ( -1.73, 8.64) | 0.192 | 9 | 79 | 9,534 |
| ***Secondary outcome*** | | | | | | | | | | | | |
| *Kidney recovery outcome* | | | | | | | | | | | | |
| Last eGFR in 90 days (ml/min/1.73m$^2$) | 74.51 | 4.11 ( -1.79, 11.73) | 0.149 | 9 | 53 | 2,225 | 84.05 | -0.85 ( -4.14, 3.84) | 0.942 | 8 | 39 | 4,863 |
| *Care process outcome* | | | | | | | | | | | | |
| Proteinuria check in 365 days (%) | 46.18 | -1.28 ( -9.42, 6.85) | 0.757 | 18 | 70 | 5,428 | 13.77 | 2.51 ( -1.36, 5.83) | 0.224 | 14 | 83 | 12,722 |
| Creatinine test in 90 days (%) | 73.13 | 1.99 ( -6.99, 9.53) | 0.763 | 11 | 96 | 3,542 | 64.09 | 2.04 ( -4.61, 6.63) | 0.725 | 9 | 59 | 7,963 |
| Blood pressure measurement in 90 days (%) | 46.25 | -1.54 ( -8.97, 5.20) | 0.602 | 18 | 98 | 6,451 | 33.45 | -0.35 ( -5.74, 4.05) | 0.735 | 11 | 76 | 10,212 |
| AKI coding in primary care in 90 days (%) | 3.13 | 5.31 ( 1.84, 8.34) | 0.002** | 24 | 64 | 13,103 | 1.60 | 2.61 ( 0.86, 3.68) | 0.002** | 28 | 63 | 73,854 |
| PPI prescription in 365 days (%) | 52.70 | 3.07 ( -3.42, 10.91) | 0.306 | 24 | 62 | 11,335 | 46.27 | -1.52 ( -6.69, 3.09) | 0.471 | 14 | 96 | 11,316 |
| NSAID prescription in 365 days (%) | 8.01 | 1.36 ( -2.97, 6.22) | 0.489 | 15 | 99 | 3,989 | 9.28 | 3.64 ( 0.69, 7.47) | 0.018* | 10 | 63 | 7,709 |
| RAASI prescription in 180 days (%) | 86.42 | -0.88 ( -8.13, 8.17) | 0.996 | 16 | 60 | 2,352 | 85.58 | -3.53 (-11.11, 3.37) | 0.295 | 14 | 75 | 2,934 |
| Diuretic prescription in 180 days (%) | 83.89 | 2.12 ( -6.18, 9.63) | 0.669 | 30 | 81 | 4,638 | 82.52 | 0.73 ( -7.02, 6.95) | 0.992 | 20 | 95 | 3,972 |

**Table S6: Effects of AKI alerts in community setting by (A) resident neighbourhood measures of deprivation (WIMD), (B) rurality (C) sex, (D) history of vascular disease and (E) diabetes.**

| | Hospital setting | | | | | | Community setting | | | | | |
|---|---|---|---|---|---|---|---|---|---|---|---|---|
| | Mean below threshold | Effect of AKI alert | | Bandwidth | | Effective sample size | Mean below threshold | Effect of AKI alert | | Bandwidth | | Effective sample size |
| | | Estimate (95% CI) | P value | below | above | | | Estimate (95% CI) | P value | below | above | |
| ***Primary outcome*** | | | | | | | | | | | | |
| Death in 365 days (%) | 29.81 | 1.31 (-3.07, 4.74) | 0.674 | 5 | 45 | 34,685 | 25.32 | 2.07 (-3.44, 6.65) | 0.534 | 7 | 70 | 9,590 |
| Emergency hospitalization or emergency department attendance in 180 days (%) | 52.98 | 0.13 (-3.82, 4.21) | 0.924 | 6 | 97 | 33,793 | 61.18 | 4.07 (-1.84, 8.27) | 0.212 | 8 | 68 | 10,592 |
| ***Secondary outcome*** | | | | | | | | | | | | |
| *Heath outcome* | | | | | | | | | | | | |
| Death in 30 days (%) | 11.66 | 0.43 (-2.33, 2.52) | 0.941 | 7 | 45 | 37,112 | 6.64 | 0.58 (-2.02, 2.30) | 0.9 | 13 | 55 | 17,598 |
| Emergency hospitalization or emergency department attendance in 30 days (%) | 26.00 | -0.35 (-3.33, 2.54) | 0.791 | 8 | 116 | 39,261 | 40.15 | 2.98 (-3.11, 7.12) | 0.442 | 8 | 48 | 11,284 |
| *Kidney recovery outcome* | | | | | | | | | | | | |
| Peak inpatient creatinine after test (micromol/L) | 106.80 | 6.45 (-0.46, 11.16) | 0.071 | 7 | 84 | 33,993 | | | | | | |
| Updated eGFR at 90 days (ml/min/1.73m$^2$) | 80.04 | -0.40 (-2.75, 2.55) | 0.941 | 5 | 40 | 23,648 | 81.51 | 0.80 (-2.48, 5.30) | 0.479 | 7 | 38 | 6,130 |
| *Care process outcome* | | | | | | | | | | | | |
| Length of inpatient stay >=7 days (%) | 47.21 | -3.09 (-8.12, 0.78) | 0.106 | 5 | 49 | 32,779 | | | | | | |
| Having an inpatient creatinine retest (%) | 70.95 | 0.80 (-3.80, 4.21) | 0.919 | 5 | 44 | 31,945 | | | | | | |
| Proteinuria check in 365 days (%) | 21.84 | 4.33 ( 0.76, 7.28) | 0.016* | 7 | 85 | 28,551 | 21.50 | 1.15 (-3.76, 5.03) | 0.777 | 10 | 73 | 10,417 |
| Creatinine test in 90 days (%) | 57.98 | 1.71 (-3.12, 5.54) | 0.584 | 6 | 73 | 28,577 | 66.29 | 3.59 (-2.67, 8.05) | 0.326 | 7 | 66 | 8,788 |
| Blood pressure measurement in 90 days (%) | 35.38 | 3.79 ( 0.58, 6.79) | 0.020* | 9 | 150 | 37,947 | 36.56 | -0.56 (-5.63, 3.42) | 0.633 | 10 | 76 | 12,225 |
| AKI coding in primary care in 90 days (%) | 3.30 | 0.72 (-0.67, 1.30) | 0.527 | 17 | 44 | 72,088 | 1.97 | 3.27 ( 1.56, 4.25) | <0.001*** | 25 | 64 | 61,535 |
| AKI coding for hospitalization (%) | 16.08 | 5.88 ( 2.22, 7.58) | <0.001*** | 9 | 29 | 36,152 | | | | | | |
| Any PPI prescription in 365 days (%) | 44.95 | 2.10 (-1.65, 5.43) | 0.295 | 8 | 130 | 32,021 | 47.80 | -1.52 (-6.28, 3.07) | 0.501 | 12 | 86 | 11,762 |
| Any NSAID prescription in 365 days (%) | 9.91 | 0.23 (-1.73, 2.53) | 0.712 | 8 | 98 | 29,016 | 8.97 | 3.19 ( 0.57, 6.56) | 0.020* | 9 | 63 | 9,215 |
| RAASI continuation in 180 days among prior users (%) | 80.93 | 2.29 (-2.09, 7.39) | 0.274 | 10 | 68 | 11,913 | 85.90 | -2.71 (-8.74, 3.39) | 0.387 | 11 | 68 | 3,779 |
| Diuretic continuation in 180 days among prior users (%) | 79.72 | 5.07 ( 0.01, 10.18) | 0.050* | 13 | 81 | 9,286 | 82.99 | 1.76 (-4.94, 6.73) | 0.764 | 18 | 84 | 5,457 |

**Table S7: Effect of AKI alert, including the MSE-optimal bandwidths[1,2,3]**

[1] Mean below threshold is calculated as the mean within the 5 micromol/L below the threshold.

[2] Effective sample size is the number of observations within the MSE-optimal bandwidth.

[3] Abbreviations: AKI = acute kidney injury

| Outcome | Robustness check | Effect of AKI alert | | | |
|---|---|---|---|---|---|
| | | Hospital setting | | Community setting | |
| | | Estimate (95% CI) | P value | Estimate (95% CI) | P value |
| Death in 365 days (%) | Main specification | 1.31 ( -3.07, 4.74) | 0.674 | 2.07 ( -3.44, 6.65) | 0.534 |
| | Quadratic fit | 0.18 ( -2.67, 2.71) | 0.989 | 1.72 ( -1.93, 5.48) | 0.348 |
| | 0.75*MSE-optimal bandwidth | 1.76 ( -3.82, 6.58) | 0.604 | 3.16 ( -3.58, 9.07) | 0.395 |
| | 1.25*MSE-optimal bandwidth | 1.22 ( -2.65, 4.04) | 0.683 | 1.88 ( -3.13, 5.61) | 0.577 |
| | 1.5*MSE-optimal bandwidth | 1.16 ( -2.44, 3.53) | 0.72 | 1.91 ( -2.99, 4.82) | 0.645 |
| | 2*MSE-optimal bandwidth | 1.78 ( -1.56, 3.34) | 0.478 | 2.84 ( -1.93, 4.59) | 0.423 |
| | Alternative index test | 3.29 ( -1.33, 7.03) | 0.182 | | |
| Emergency hospitalization or emergency department attendance in 180 days (%) | Main specification | 0.13 ( -3.82, 4.21) | 0.924 | 4.07 ( -1.84, 8.27) | 0.212 |
| | Quadratic fit | 0.95 ( -1.62, 3.76) | 0.435 | 3.06 ( -1.31, 6.45) | 0.195 |
| | 0.75*MSE-optimal bandwidth | -0.25 ( -5.06, 4.67) | 0.938 | 3.29 ( -3.54, 8.84) | 0.401 |
| | 1.25*MSE-optimal bandwidth | 0.53 ( -2.79, 3.97) | 0.731 | 4.42 ( -0.81, 7.91) | 0.111 |
| | 1.5*MSE-optimal bandwidth | 0.25 ( -2.76, 3.28) | 0.864 | 4.92 ( 0.15, 7.90) | 0.042* |
| | 2*MSE-optimal bandwidth | 0.23 ( -2.37, 2.61) | 0.924 | 5.79 ( 1.44, 7.91) | 0.005** |
| | Alternative index test | -0.68 ( -4.68, 3.53) | 0.784 | | |
| Death in 30 days (%) | Main specification | 0.43 ( -2.33, 2.52) | 0.941 | 0.58 ( -2.02, 2.30) | 0.9 |
| | Quadratic fit | 0.26 ( -1.75, 2.08) | 0.868 | 0.11 ( -2.09, 2.08) | 0.993 |
| | 0.75*MSE-optimal bandwidth | 0.56 ( -2.67, 3.40) | 0.816 | 0.48 ( -2.46, 2.61) | 0.954 |
| | 1.25*MSE-optimal bandwidth | 0.66 ( -1.91, 2.27) | 0.866 | 0.80 ( -1.65, 2.17) | 0.787 |
| | 1.5*MSE-optimal bandwidth | 0.95 ( -1.53, 2.20) | 0.723 | 1.12 ( -1.26, 2.21) | 0.594 |
| | 2*MSE-optimal bandwidth | 1.24 ( -1.13, 1.96) | 0.595 | 1.65 ( -0.82, 2.20) | 0.37 |
| | Alternative index test | 1.75 ( -1.74, 4.42) | 0.393 | | |
| Emergency hospitalization or emergency department attendance in 30 days (%) | Main specification | -0.35 ( -3.33, 2.54) | 0.791 | 2.98 ( -3.11, 7.12) | 0.442 |
| | Quadratic fit | -0.39 ( -2.47, 1.73) | 0.73 | 2.12 ( -2.59, 5.89) | 0.447 |
| | 0.75*MSE-optimal bandwidth | -0.81 ( -4.44, 2.71) | 0.635 | 1.86 ( -5.00, 7.46) | 0.699 |
| | 1.25*MSE-optimal bandwidth | -0.36 ( -2.94, 2.11) | 0.745 | 3.26 ( -2.39, 6.47) | 0.366 |
| | 1.5*MSE-optimal bandwidth | -0.23 ( -2.57, 1.90) | 0.77 | 3.86 ( -1.41, 6.45) | 0.209 |
| | 2*MSE-optimal bandwidth | 0.00 ( -2.00, 1.69) | 0.865 | 4.96 ( 0.14, 6.72) | 0.041* |
| | Alternative index test | 0.44 ( -2.75, 3.73) | 0.767 | | |
| Having an inpatient creatinine retest (%) | Main specification | 0.80 ( -3.80, 4.21) | 0.919 | | |
| | Quadratic fit | 1.88 ( -1.01, 4.49) | 0.216 | | |
| | 0.75*MSE-optimal bandwidth | 0.31 ( -5.31, 4.67) | 0.9 | | |
| | 1.25*MSE-optimal bandwidth | 1.50 ( -2.42, 4.20) | 0.599 | | |
| | 1.5*MSE-optimal bandwidth | 3.13 ( -0.42, 5.38) | 0.093 | | |
| | 2*MSE-optimal bandwidth | 4.76 ( 1.41, 6.22) | 0.002** | | |
| | Alternative index test | 5.13 ( 1.55, 7.68) | 0.003** | | |
| Updated eGFR at 90 days (ml/min/1.73m2) | Main specification | -0.40 ( -2.75, 2.55) | 0.941 | 0.80 ( -2.48, 5.30) | 0.479 |
| | Quadratic fit | 0.10 ( -1.57, 2.12) | 0.772 | 1.01 ( -1.93, 4.26) | 0.46 |
| | 0.75*MSE-optimal bandwidth | 0.65 ( -2.66, 4.40) | 0.629 | 0.96 ( -3.32, 6.35) | 0.539 |
| | 1.25*MSE-optimal bandwidth | -1.20 ( -3.10, 1.45) | 0.478 | 0.07 ( -2.57, 4.17) | 0.643 |
| | 1.5*MSE-optimal bandwidth | -1.71 ( -3.27, 0.81) | 0.238 | -0.76 ( -2.92, 3.13) | 0.946 |
| | 2*MSE-optimal bandwidth | -2.50 ( -3.41, -0.07) | 0.042* | -2.42 ( -3.63, 1.39) | 0.381 |
| | Alternative index test | -1.35 ( -3.59, 1.70) | 0.485 | | |

| | | | | | |
|---|---|---|---|---|---|
| Peak inpatient creatinine after test (micromol/L) | Main specification | 6.45 ( -0.46, 11.16) | 0.071 | | |
| | Quadratic fit | 2.88 ( -2.12, 7.19) | 0.285 | | |
| | 0.75*MSE-optimal bandwidth | 6.75 ( -1.23, 13.34) | 0.103 | | |
| | 1.25*MSE-optimal bandwidth | 6.01 ( -0.35, 9.85) | 0.068 | | |
| | 1.5*MSE-optimal bandwidth | 6.16 ( -0.06, 9.10) | 0.053 | | |
| | 2*MSE-optimal bandwidth | 6.86 ( 0.62, 8.36) | 0.023* | | |
| | Alternative index test | 7.50 ( 0.53, 11.95) | 0.032* | | |
| Diuretic continuation in 180 days among prior users (%) | Main specification | 5.07 ( 0.01, 10.18) | 0.050* | 1.76 ( -4.94, 6.73) | 0.764 |
| | Quadratic fit | 4.22 ( -0.84, 9.25) | 0.102 | -0.88 ( -8.00, 4.84) | 0.629 |
| | 0.75*MSE-optimal bandwidth | 4.45 ( -1.75, 10.44) | 0.162 | 2.02 ( -5.54, 8.11) | 0.712 |
| | 1.25*MSE-optimal bandwidth | 5.05 ( 0.69, 9.53) | 0.024* | 1.24 ( -4.48, 5.80) | 0.802 |
| | 1.5*MSE-optimal bandwidth | 4.14 ( 0.28, 8.14) | 0.036* | 1.13 ( -3.94, 5.38) | 0.761 |
| | 2*MSE-optimal bandwidth | 2.66 ( -0.52, 5.97) | 0.1 | 1.91 ( -2.29, 5.91) | 0.387 |
| | Inclusion of non-survivors | 4.95 ( 0.51, 9.85) | 0.030* | 0.25 ( -5.07, 5.49) | 0.938 |
| | Alternative index test | 5.53 ( -0.02, 11.27) | 0.051 | | |
| Any NSAID prescription in 365 days (%) | Main specification | 0.23 ( -1.73, 2.53) | 0.712 | 3.19 ( 0.57, 6.56) | 0.020* |
| | Quadratic fit | -0.24 ( -2.19, 1.12) | 0.525 | 3.22 ( 0.59, 5.99) | 0.017* |
| | 0.75*MSE-optimal bandwidth | 1.02 ( -1.42, 3.72) | 0.381 | 4.20 ( 0.86, 7.99) | 0.015* |
| | 1.25*MSE-optimal bandwidth | -0.01 ( -1.63, 2.07) | 0.817 | 2.87 ( 0.70, 5.99) | 0.013* |
| | 1.5*MSE-optimal bandwidth | -0.16 ( -1.55, 1.75) | 0.906 | 2.63 ( 0.76, 5.55) | 0.010** |
| | 2*MSE-optimal bandwidth | -0.55 ( -1.65, 1.09) | 0.691 | 1.86 ( 0.41, 4.53) | 0.019* |
| | Inclusion of non-survivors | 0.26 ( -1.47, 2.25) | 0.68 | 2.24 ( 0.01, 5.10) | 0.050* |
| | Alternative index test | -0.48 ( -2.49, 1.73) | 0.725 | | |
| Any PPI prescription in 365 days (%) | Main specification | 2.10 ( -1.65, 5.43) | 0.295 | -1.52 ( -6.28, 3.07) | 0.501 |
| | Quadratic fit | 0.23 ( -2.74, 3.23) | 0.872 | -1.07 ( -5.49, 3.25) | 0.615 |
| | 0.75*MSE-optimal bandwidth | 4.03 ( -0.35, 8.21) | 0.072 | -2.23 ( -7.77, 3.45) | 0.451 |
| | 1.25*MSE-optimal bandwidth | 1.32 ( -2.07, 4.04) | 0.528 | -0.78 ( -5.11, 3.07) | 0.624 |
| | 1.5*MSE-optimal bandwidth | 1.05 ( -2.06, 3.36) | 0.638 | -0.84 ( -4.79, 2.56) | 0.554 |
| | 2*MSE-optimal bandwidth | 1.00 ( -1.67, 2.82) | 0.615 | -0.59 ( -4.02, 2.23) | 0.573 |
| | Inclusion of non-survivors | 2.64 ( -0.67, 5.79) | 0.121 | -1.66 ( -5.29, 2.31) | 0.443 |
| | Alternative index test | -1.18 ( -5.32, 2.53) | 0.486 | | |
| RAASI continuation in 180 days among prior users (%) | Main specification | 2.29 ( -2.09, 7.39) | 0.274 | -2.71 ( -8.74, 3.39) | 0.387 |
| | Quadratic fit | 0.58 ( -3.34, 4.55) | 0.764 | -1.85 ( -7.19, 3.21) | 0.454 |
| | 0.75*MSE-optimal bandwidth | 4.25 ( -1.21, 10.27) | 0.122 | -3.66 (-10.99, 3.40) | 0.301 |
| | 1.25*MSE-optimal bandwidth | 1.00 ( -2.58, 5.59) | 0.47 | -2.78 ( -7.80, 2.88) | 0.366 |
| | 1.5*MSE-optimal bandwidth | 0.25 ( -2.71, 4.55) | 0.62 | -2.67 ( -6.94, 2.74) | 0.395 |
| | 2*MSE-optimal bandwidth | -0.82 ( -2.96, 3.02) | 0.983 | -1.57 ( -4.78, 3.54) | 0.77 |
| | Inclusion of non-survivors | 2.37 ( -2.37, 7.83) | 0.294 | -6.01 (-12.62, 0.35) | 0.064 |
| | Alternative index test | 2.41 ( -2.53, 8.07) | 0.305 | | |
| AKI coding for hospitalization (%) | Main specification | 5.88 ( 2.22, 7.58) | <0.001*** | | |
| | Quadratic fit | 5.87 ( 3.07, 7.20) | <0.001*** | | |
| | 0.75*MSE-optimal bandwidth | 4.71 ( 0.78, 7.27) | 0.015* | | |
| | 1.25*MSE-optimal bandwidth | 6.67 ( 3.14, 7.80) | <0.001*** | | |
| | 1.5*MSE-optimal bandwidth | 7.53 ( 4.07, 8.22) | <0.001*** | | |
| | 2*MSE-optimal bandwidth | 9.38 ( 5.96, 9.44) | <0.001*** | | |
| | Alternative index test | 6.23 ( 2.50, 7.28) | <0.001*** | | |

| | | | | | |
|---|---|---|---|---|---|
| AKI coding in primary care in 90 days (%) | Main specification | 0.72 ( -0.67, 1.30) | 0.527 | 3.27 ( 1.56, 4.25) | <0.001*** |
| | Quadratic fit | 0.67 ( -0.44, 1.32) | 0.329 | 3.06 ( 1.38, 4.28) | <0.001*** |
| | 0.75*MSE-optimal bandwidth | 0.38 ( -1.13, 1.22) | 0.935 | 3.10 ( 1.45, 4.47) | <0.001*** |
| | 1.25*MSE-optimal bandwidth | 1.22 ( -0.12, 1.61) | 0.091 | 3.46 ( 1.86, 4.33) | <0.001*** |
| | 1.5*MSE-optimal bandwidth | 1.63 ( 0.30, 1.86) | 0.006** | 3.69 ( 2.15, 4.50) | <0.001*** |
| | 2*MSE-optimal bandwidth | 2.47 ( 1.11, 2.44) | <0.001*** | 4.26 ( 2.64, 4.84) | <0.001*** |
| | Inclusion of non-survivors | 0.75 ( -0.59, 1.32) | 0.456 | 3.35 ( 1.78, 4.19) | <0.001*** |
| | Alternative index test | 0.99 ( -0.48, 1.47) | 0.322 | | |
| Blood pressure measurement in 90 days (%) | Main specification | 3.79 ( 0.58, 6.79) | 0.020* | -0.56 ( -5.63, 3.42) | 0.633 |
| | Quadratic fit | 2.92 ( 0.13, 5.72) | 0.040* | -0.97 ( -5.38, 3.01) | 0.579 |
| | 0.75*MSE-optimal bandwidth | 4.40 ( 0.58, 8.09) | 0.024* | -1.03 ( -6.87, 4.00) | 0.605 |
| | 1.25*MSE-optimal bandwidth | 3.26 ( 0.43, 5.79) | 0.023* | -0.06 ( -4.68, 3.24) | 0.721 |
| | 1.5*MSE-optimal bandwidth | 3.04 ( 0.47, 5.22) | 0.019* | 0.26 ( -3.96, 3.11) | 0.813 |
| | 2*MSE-optimal bandwidth | 2.94 ( 0.76, 4.69) | 0.007** | 1.07 ( -2.60, 3.40) | 0.795 |
| | Inclusion of non-survivors | 4.21 ( 1.20, 7.28) | 0.006** | -0.85 ( -5.14, 2.90) | 0.584 |
| | Alternative index test | 3.13 ( -0.60, 6.77) | 0.101 | | |
| Creatinine test in 90 days (%) | Main specification | 1.71 ( -3.12, 5.54) | 0.584 | 3.59 ( -2.67, 8.05) | 0.326 |
| | Quadratic fit | 0.14 ( -3.12, 2.90) | 0.944 | 0.26 ( -4.37, 4.01) | 0.933 |
| | 0.75*MSE-optimal bandwidth | 1.76 ( -4.09, 6.72) | 0.633 | 4.48 ( -2.72, 10.33) | 0.253 |
| | 1.25*MSE-optimal bandwidth | 1.95 ( -2.35, 5.14) | 0.466 | 3.07 ( -2.65, 6.65) | 0.398 |
| | 1.5*MSE-optimal bandwidth | 1.82 ( -2.10, 4.46) | 0.48 | 2.47 ( -2.77, 5.53) | 0.514 |
| | 2*MSE-optimal bandwidth | 2.22 ( -1.28, 4.13) | 0.302 | 3.01 ( -1.60, 5.33) | 0.291 |
| | Inclusion of non-survivors | 1.57 ( -2.92, 5.20) | 0.581 | 3.26 ( -2.07, 7.90) | 0.251 |
| | Alternative index test | 2.62 ( -2.44, 6.84) | 0.353 | | |
| Length of inpatient stay >=7 days (%) | Main specification | -3.09 ( -8.12, 0.78) | 0.106 | | |
| | Quadratic fit | -0.92 ( -4.20, 2.10) | 0.515 | | |
| | 0.75*MSE-optimal bandwidth | -2.92 ( -9.18, 2.30) | 0.24 | | |
| | 1.25*MSE-optimal bandwidth | -1.67 ( -6.07, 1.40) | 0.221 | | |
| | 1.5*MSE-optimal bandwidth | -0.28 ( -4.35, 2.29) | 0.543 | | |
| | 2*MSE-optimal bandwidth | 1.41 ( -2.42, 3.05) | 0.822 | | |
| | Inclusion of non-survivors | -3.18 ( -7.95, 0.61) | 0.093 | | |
| | Alternative index test | 0.90 ( -3.86, 4.55) | 0.872 | | |
| Proteinuria check in 365 days (%) | Main specification | 4.33 ( 0.76, 7.28) | 0.016* | 1.15 ( -3.76, 5.03) | 0.777 |
| | Quadratic fit | 2.33 ( -0.29, 4.63) | 0.084 | 1.31 ( -2.66, 4.73) | 0.582 |
| | 0.75*MSE-optimal bandwidth | 4.02 ( -0.14, 7.69) | 0.059 | -0.52 ( -6.30, 4.23) | 0.701 |
| | 1.25*MSE-optimal bandwidth | 4.34 ( 1.10, 6.80) | 0.007** | 2.24 ( -2.16, 5.49) | 0.394 |
| | 1.5*MSE-optimal bandwidth | 4.23 ( 1.31, 6.35) | 0.003** | 2.67 ( -1.37, 5.47) | 0.24 |
| | 2*MSE-optimal bandwidth | 3.56 ( 1.05, 5.23) | 0.003** | 3.08 ( -0.61, 5.20) | 0.121 |
| | Inclusion of non-survivors | 3.73 ( 0.94, 6.03) | 0.007** | 1.55 ( -1.87, 4.48) | 0.42 |
| | Alternative index test | 3.50 ( -0.27, 6.53) | 0.071 | | |

**Table S8: Robustness checks for the effects of AKI alert in hospital and community setting[1,2,3]**

[1]Alternative index test” refers to the test with the highest reading in the first two days of inpatient stay, and inpatient stays of less than two days are excluded from this specific robustness check.

[2] Abbreviations: AKI = acute kidney injury
